# A Health Economic Assessment of Relaxation of COVID-19 Control in England 2021

**DOI:** 10.1101/2025.10.24.25338659

**Authors:** A.J. Cooper, Matt J. Keeling

**Affiliations:** SBIDER (Systems Biology and Infectious Disease Epidemiology Research) Institute, University of Warwick. Coventry. CV4 7AL; Mathematics Institute, University of Warwick. Coventry. CV4 7AL; School of Life Sciences, University of Warwick. Coventry. CV4 7AL

## Abstract

The SARS-CoV-2 pandemic caused devastating health, social and economic harms. Globally, during the first year of this pandemic, controls were primarily focused on containing the outbreak and minimising infection, disease and health consequences. However, during the second year (2021) with vaccines being deployed, there was a recognition that mitigation measures needed to consider both infection control and socio-economic priorities, inevitably causing a tension between how these conflicting elements are balanced. In England, this was epitomised by the Roadmap out of Lockdown, which through 4 planned steps reduced controls from the January 2021 lockdown to no restrictions on social interactions. Here, we adopt a health-economic framework to consider a range of alternative approaches to the relaxation of controls, allowing us to quantitatively assess health losses (measured in Quality Adjusted Life Years, QALYs) and economic consequences (measured in terms of Gross Domestic Product, GDP) in a unified framework. Using the UK Treasury framework of valuing one QALY at £70,000, we find that the implemented Roadmap performs extremely well, only being consistently outperformed by a similar strategy that starts all steps one week earlier. This work highlights the power of a holistic framework combining epidemiology and economics, and suggests its utility in future outbreaks.

## 1 Introduction

Social restrictions are a highly effective means of controlling an outbreak; by reducing the interactions between all members of society, the critical mixing between infectious and susceptible individuals is also reduced. While this method of control is robust and does not require a detailed understanding of the pathogen, it is an untargeted approach that has broad implications for society. In the UK and elsewhere, restrictions on social mixing had a considerable impact on the transmission of SARS-CoV-2 [1], however it was not viable for these to remain in place indefinitely. After the third UK lockdown (beginning on 4th January 2021), and with vaccination predicted to generate population-level protection, the government wished to ease restrictions (in a series of well-defined steps) without the risk of overwhelming the health services. In consultation with scientific advisors and the Treasury, the government published the Roadmap out of Lockdown on the 21st February 2021 [2], as a plan for gradual relaxation of restrictions in England. (The Devolved Administrations of Scotland, Wales and Northern Ireland had similar, but independent plans).

The roadmap was built around the projected vaccine rollout [3]: by the middle of Februrary, everyone in the top four priority groups (which included residents and staff in care homes for older adults, all those over 70 years of age, frontline health and social care workers and extremely clinically vulnerable individuals) would have been offered at least one dose of the vaccine. The subsequent aims were for first doses to be offered to all those in groups 5-9 (all those over 50 years of age and those in an at-risk group over 16 years of age) by 15th April and then for all adults to have been offered a first dose by the end of July. This increasing level of protection against infection and disease formed a cornerstone of public health planning, and (if protection was long lasting) would eventually eliminate the need for other forms of control [4].

The Government stressed that the relaxation of restrictions must be irreversible, and in order to ensure this, stated that moving to the next step of the roadmap would be led only by data (not fixed dates) and would be subject to four tests:

(1) The vaccine deployment program continued successfully.
(2) Evidence showed that vaccines were sufficiently effective in reducing hospitalisations and death.
(3) Infection rates did not risk a surge in hospitalisations which would put unsustainable pressure on the NHS.
(4) Any risk assessment was not fundamentally changed by new variants of concern. There were at least five weeks between each step, allowing four weeks for the impact of the previous step to be detected in the data and one week to give the public notice before any changes. This timescale aimed to ensure that the impact of each step could be properly assessed, and reduce the likelihood of having to reverse course and reimpose any restrictions. Throughout, modelling and statistical analysis of the data provided critical input on defining the time course of relaxation, and providing an assessment that tests 3 and 4 (above) were being met and would continue to be met following the next relaxation step.

The enacted roadmap consisted of four basic steps, each of which lifted a set of social mixing restrictions, allowing the country to transition from full lockdown to an absence of social controls [2]:

**Lockdown** (4th January – 7th March 2021). Stay at home order: Shopping for basic necessities only; non-essential retail, hospitality and personal care services closed. Work from home where possible. Can leave home for exercise (with household, support bubble or one other person), to meet support bubble, or to seek medical care. Remote learning in schools except key workers and vulnerable. Clinically extremely vulnerable advised to shield.

**Step 1a** (8th March 2021). Schools reopen, with twice-weekly lateral flow testing of staff and pupils.

**Step 1b** (29th March 2021). Meeting outdoors with 6 people or 2 households allowed. Outdoor sports facilities can reopen.

**Step 2** (not before 12th April 2021). Non-essential retail, personal care and public buildings can reopen. Outdoor table service in hospitality venues.

**Step 3** (not before 17th May 2021). Meeting outdoors with up to 30 people, and indoors with 6 people or 2 households allowed. Most businesses can reopen, including indoor hospitality.

**Step 4** (not before 21st June 2021). All legal limits on social contact removed.

Three teams from UK universities provided modelling input, through SPI-M-O and SAGE, to support the roadmap: Imperial College; London School of Hygiene and Tropical Medicine; and University of Warwick. The first roadmap document was concerned with the general timescale of relaxation, which was considered a balance between sufficient immunity from vaccination and greater freedom for increased social mixing [5]. Subsequent roadmaps addressed the epidemiological trends and likely impacts of the subsequent relaxation steps, providing confidence to the government that none of the step changes would overwhelm healthcare resources [6–9]. In response to uncertainty around the emergence of the Delta variant, a one month delay was enacted in June which postponed the implementation of the fourth and final step of the roadmap until 19th July 2021.

The roadmap was completed in 2021 with the stated aim of opening up the country without overwhelming any health services. Throughout the course of the relaxation the economy grew steadily (in terms of GDP) with quarterly increases of 7.3%, 1.7% and 1.5% in the last three quarters of 2021. The impact of the roadmap steps can be judged in terms direct health impacts, using a health-economic analysis (considering costs to the NHS and loss of quality adjusted life years (QALYs) following infection, hospitalisation or death) and an economic analysis (considering the impact of these steps on the gross domestic product (GDP) of England). Through a retrospective model-based analysis of 2021 (from 1st March to 30th November – around the time when the Omicron variant emerged in the UK), we quantify the impact of the realised timeline and compare this to a series of alternative scenarios. The adopted roadmap for relaxation can be contrasted with calls at the time for a more accelerated relaxation in which all controls were lifted by the end of April 2021 [10], and with a more cautious approach in which England remained under some restrictions for far longer [10]. These two scenarios, together with a series of others where the relaxation steps are shorter, longer, implemented earlier or later are also considered. In addition, the effectiveness of the one month delay introduced in response to uncertainty around the emergence of the Delta variant is also considered.

To compare alternative scenarios requires a transmission model that can translate the social mixing within each step into an epidemiological forecast – this non-linear process is further complicated by the changing level of vaccine protection over time. The Warwick age-structured SARS-CoV-2 transmission model provides the necessary framework. This model was matched to a variety of epidemic data from England throughout the pandemic [11]; it was used to inform the Roadmap steps [12–16] and these projections have retrospectively been shown to compare well with the true number of hospital admissions [17]. For each alternative scenario, the output from the model was combined with GDP estimates to calculate the total losses, as well as an assessment of whether test 3 (unsustainable pressure on the NHS) held.

Our combined approach, which considers both the health economics of COVID-19 disease and the national economy, provides an effective method of robustly and quantitatively comparing different approaches to opening up the economy. As such, we obtain an objective means of balancing economic and public health considerations; the only arbitrary choice is the value placed on one year of healthy life.

## 2 Methods

We begin by considering the suite of alternative scenarios that will be modelled throughout this paper, we then outline the transmission model, the health economic framework that is used to translate the epidemiological model output into an economic value, and finally the economic assessment that considers how changes to the roadmap would likely scale the GDP of England. Throughout our analysis we are interested in the nine-month period from 1st March 2021 to 30th November 2021 – from the later stages of the 2021 lockdown to the rise of Omicron.

### 2.1 Roadmap Scenarios

Here we consider nine alternative scenarios based on the four steps defined in the initial roadmap documentation [2]. These alternatives range in the speed with which the UK would progress from full lockdown to Step 4 where all limits on social contacts are removed (Figure 1).

**Figure 1:**
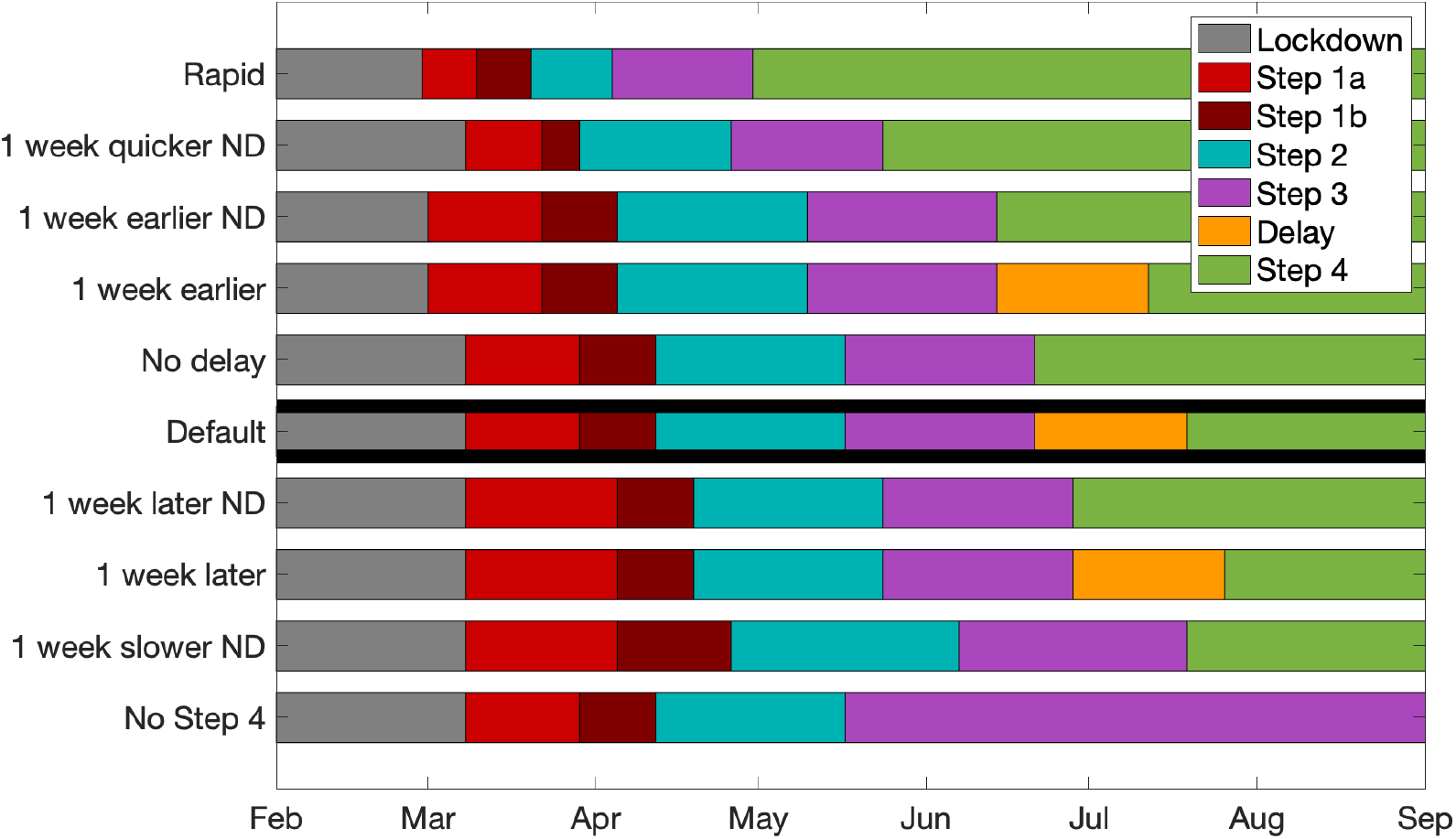
Timelines for the default Roadmap and alternative scenarios. These ten strategies are ordered in terms of when step 3 is completed; throughout ND denotes no delay to the final step.

Some groups advocated for a quicker return of economic activity once the most vulnerable were protected by vaccination, and there was pressure for a timeline based on firm dates: some schools to open in February, pubs and restaurants to open by Easter and all restrictions to be removed by the end of April when most vulnerable groups should have been vaccinated. This becomes the fastest of our alternative roadmap timelines, denoted by the label *Rapid*.

A *One week quicker* strategy involves each of the implemented steps being a week shorter so that all restrictions are lifted by 24th May. A *One week earlier* scenario shifts the whole roadmap a week earlier, although maintains the duration of each step. We examine this strategy with (*One week earlier*) and without (*One week earlier ND*) a 4-week delay caused by Delta variant uncertainty. The *No delay* strategy maintains the observed timings of steps 1 to 3, but does not implement the delay to Step 4, allowing it to occur on 21st June 2021. These five strategies are all associated with a more rapid relaxation of controls than was observed.

Slower relaxation scenarios are also considered forming counterparts to the faster strategies. A one week later strategy enacts Steps 1b to 4 one week later than the default case; this is implemented with (*One week later*) and without (*One week later ND*) the Delta delay. A *One week slower* strategy stretches each step by a week, providing more time to assess the changes due to the previous step. Finally, the most cautious strategy, requiring more stringent restrictions to be in place for a prolonged period of time, is represented by a timeline where the final step is not implemented and the country remains under Step 3 restrictions for the duration of the analysis, described as *No Step 4*.

We note that the issue of whether Step 4 is delayed by the uncertainties of Delta variant is only relevant for a limited number of scenarios (1 week earlier, default and 1 week later) where the timing of Step 4 is in June 2021.

### 2.2 Transmission model

Here we briefly describe the mathematical foundations of the Warwick age-structured SARS-CoV-2 transmission model [4, 11, 17–20] which was developed to generate policy advice throughout the progression of the UK pandemic. The model stems from the deterministic SEIR (Susceptible, Exposed, Infectious, Recovered) framework and is formulated with twenty-one five-year age groups (0-4, 5-9, …, 95-99, 100+). It captures the restrictions on social interaction but maintains transmission within a household.

In the initial waves of infection the main driver of infection dynamics within the model was the level of precautionary behaviour; this captures the scale of population mixing and transmission outside the household (where a precautionary behaviour value of zero corresponds to pre-pandemic levels of social mixing, and greater restrictions on social interaction are captured by precautionary behaviour between zero and one). Precautionary behaviour was inferred as a slowly varying parameter throughout the pandemic to capture the gradual change in human behaviour, with punctuated larger changes whenever new restrictions were imposed. The model was extensively modified during the pandemic to account for the emergence of new variants (Alpha, Delta, Omicron) and the introduction of vaccination. These modifications all increased the dimensionality of the model through the inclusion of additional classes for each variant and further segregation of the population by vaccination status.

Each of the default roadmap relaxation steps gives rise to a decrease in level of precautionary behaviour; we assume this level of precautionary behaviour and hence social mixing remains applicable for the same step in the alternative roadmaps. The only exception to this rule is the decrease in precautionary behaviour due to the 2020 UEFA European Football Championships, and the subsequent decrease in social mixing caused by the pingdemic (attributed to the surge in cases following the football together with the emergence of the Delta variant); these two events are assumed to occur at fixed dates. An-other key factor is the assumption that the timeline for the vaccine rollout and its uptake in all age groups is preserved. Values for the levels of vaccine protection against infection, symptomatic disease, requiring hospital admission, against death and the reduction in onward transmission for different vaccines and for different variants used in the model are given in the Supplementary Material. We also assume that the Delta variant arrives at the same time and with the same magnitude, although its initial growth depends on the chosen scenario.

Key values of public health interest that are outputs from the model are daily levels of symptomatic infection, hospital admissions, hospital occupancy, admissions to critical care and number of daily deaths. All of these are generated for each of the twenty one age groups and are proportional to the incidence of infection (often with some delay). These are the metrics required for the health-economic framework.

### 2.3 Health-economic framework

The health impacts of each scenario can be expressed in terms of the number of hospitalisations and deaths predicted in each age group. Given the stated aim that infection rates should not put unsustainable pressure on the NHS, the total level of hospital occupancy is also an important metric. The health costs associated with each scenario can be divided into direct hospitalisation costs (costs associated with treatment and care in hospital and critical care wards) and costs associated with losses to Quality Adjusted Life Years (QALYs) including deaths.

Hospital episode costs for England have been obtained from the NHS 2021/22 National Cost Collection Data Publication [21] using the appropriate Healthcare Resource Group (HRG) codes, and the average number of Finished Consultant Episodes (FCEs) per admission was obtained from NHS Digital, Hospital Episode Statistics (HES), England 2021-22 [22]. This gave a weighted average cost for each hospitalisation of £5267 for adults and £3671 for children. For critical care, the weighted average cost per day was determined to be £2144 for adults and £2371 for children.

Since the epidemic model divides the population into twenty-one five-year age groups (with age group labelled by *a*) it was considered appropriate to determine age-varying hospitalisation costs. Length of stay data for COVID-19 [23] and length of stay in critical care data [24] were used to determine age-varying admission costs (*C*^*H*^(*a*)) and critical care costs (*C*^*I*^(*a*)); in general these costs increase with age due to the longer average length of stay (see Supplementary Material).

The costs associated with quality adjusted life year (QALY) losses depend on the disease severity and are proportional to the associated loss of QALYs due to symptomatic infection (*S*), hospitalisation (*H*), ICU or critical care admission (*I*) and death (*D*). We also include losses associated with Long COVID symptoms (*L*). QALY losses associated with COVID-19 deaths are age-dependent and were sourced from the average life expectancy in the UK for 2020-2022 [25], with adjustments for age– and sex-specific QALY population norms based on the EQ-5D-3L for the UK, and also include the standard UK discounting rate of 3.5%. All other QALY losses are assumed to be independent of age, and discounting is ignored given the relatively short duration.

The overall cost of COVID-related health impacts can then be expressed as

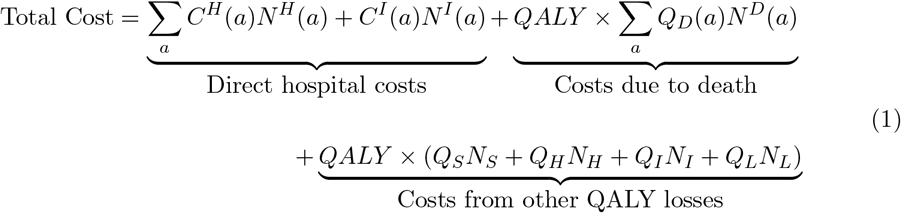

where *a* denotes age group, *N*^*H*^(*a*) or *N*^*I*^(*a*) are the number of admissions (to hospital or intensive care) in that age group, *Q*_*X*_ denotes the loss of QALYs associated with outcome *X, N*_*X*_ is the total number with outcome *X* (hence *N*_*H*_ = ∑_a_ *N*^*H*^(*a*)) and *QALY* is the monetary value assigned to one quality adjusted life year.

QALY losses due to premature death range from 23 for children under 5 to less than 3 for those over 90 years of age, with the full age profile given in the Supplementary Material. Cost calculations use the values *Q*_*S*_ = 0.008, *Q*_*H*_ = 0.0201, *Q*_*I*_ = 0.15 and *Q*_*L*_ = 0.034, with the assumption that 10% of symptomatic cases give rise to symptoms for more than 12 weeks [26]. Alternative assumptions for QALY losses are explored in the Supplementary Material, but have no qualitative impact on our results.

Estimates for indirect costs, for example those associated with delayed health care, mental health problems connected to social restrictions or productivity losses due to COVID-19 infection, hospitalisation or death have not been included, nor the cost of Long COVID to the health service. Each of these would raise the average economic burden associated with each case, and hence favour strategies that minimise infection.

This retrospective analysis is not a standard cost-effectiveness calculation [27] as we are comparing the cost of national restrictions, rather than considering the cost-effectiveness of a medical intervention. As such, we considered two quantities for the monetary value of a QALY. Firstly costs were calculated assuming one QALY is valued at £30,000, which is the value used in cost-effectiveness calculations for NICE [28] and therefore reflects usual practise for health interventions. Secondly, we use the Treasury estimate of £70,000 as the willingness to pay value for a QALY as given in The Treasury Green Book (2022) based on 2020/21 costs [29], which is considered more appropriate for national-scale economic decisions. This approach of considering two QALY values was used by Welsh Government COVID-19 TAG Policy Modelling Group during the pandemic [30].

### 2.4 Economic framework

The gross domestic product (GDP) of England over the time period of interest is taken as our measure of economic gain to set again the health losses. The adjusted UK monthly GDP data for 2021 [31] is extrapolated to give daily values for England (by fitting a spline to the cumulative data), and rescaled by the 2021 annual GDP of England (£2.409815 trillion) to convert into true monetary values. The resulting daily GDP is averaged over each step of the default roadmap to obtain an associated GDP. For alternative roadmap scenarios, this allows us to estimate the total GDP over the period, by accounting for the time in each step. As expected faster relaxation scenarios are associated with higher total GDP.

This approach does not account for any change in behaviour (and therefore economic spending) that may occur in response to the rise or fall in cases, hospitalisations and deaths, but provides a simple economic projection to compare with the health economic burden. Estimated monthly GDP for the period of interest is shown for each of the roadmap scenarios in the Supplementary Material.

## 3 Results

We begin with data for the default case of the actual roadmap and evolution of the epidemic in 2021 together with calculated health costs and monthly GDP (all for England). The time period of interest is taken to be 1st March 2021, just before the first of the relaxation steps was taken (Step 1 – 8th March) through to 30th November 2021 (just as the Omicron variant was detected in the UK). Key values of interest are shown in Figure 2. In the period of interest, our transmission model predicts a total of 130,908 (95% PI: 128,746 – 132,753) hospital admissions and 17,430 (95% PI: 16,790 – 17,956) deaths. These numbers differ slightly from the reported values (126,761 hospital admissions and 17,606 deaths), but are used to give a more natural correspondence with the alternative scenarios which can only be explored through simulation, and because the model provides the fine-scale age-structure required for the health economic analysis. The total health costs are estimated to be £5.44bn (95% PI: £5.24bn – £5.78bn) based on one QALY valued at £30,000, or £11.29bn (95% PI: £10.84bn – £12.01bn) at £70,000 per QALY, while the total GDP is £1.62tn. We note that the total health costs are dominated by the cost of QALY losses due to premature death, which comprises approximately 70% of the total health cost.

**Figure 2:**
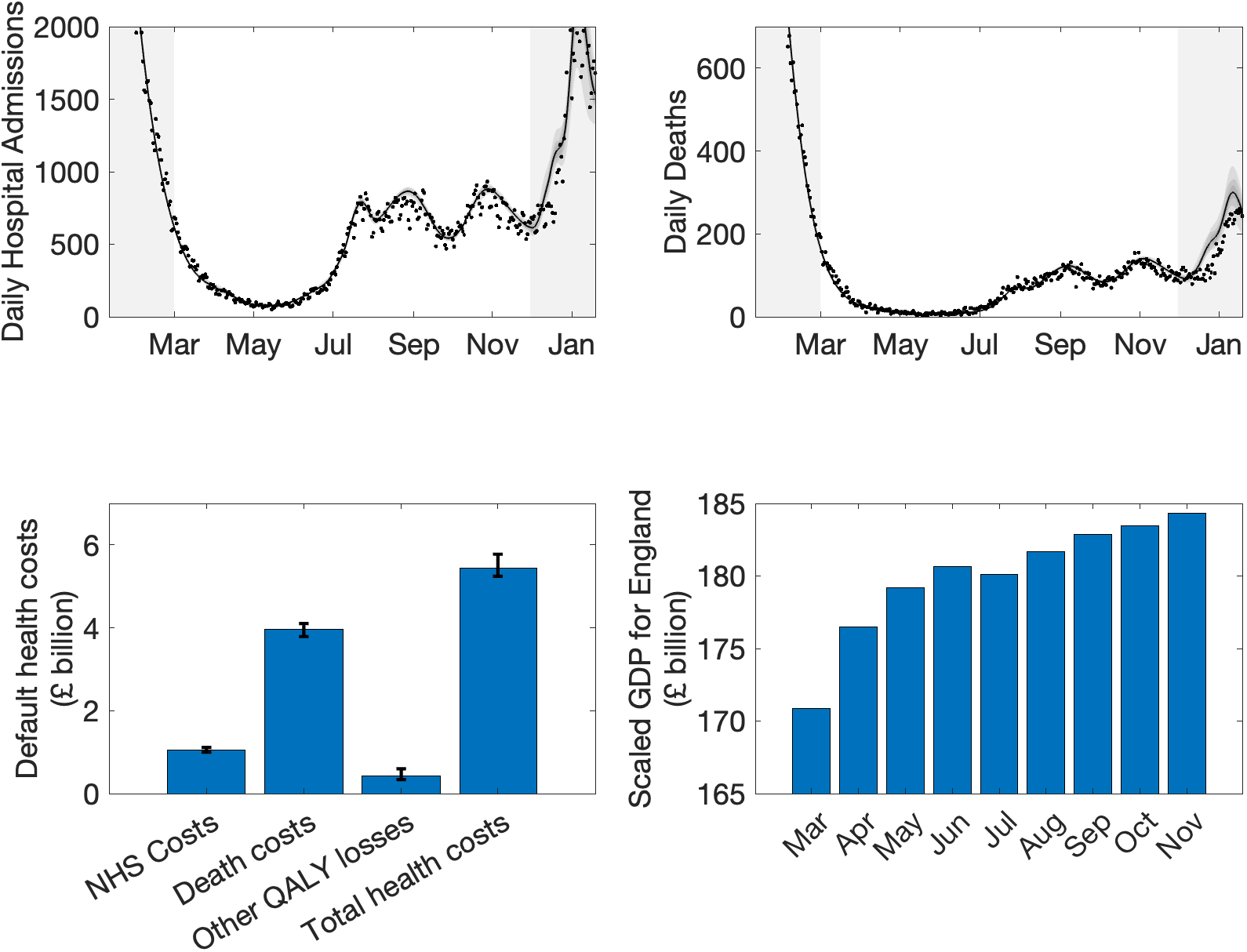
Values of interest for the default scenario. Upper panels: Daily hospital admissions and deaths in England, comparing observed outcomes (black dots) with the default model projection (black lines show mean values and shaded regions are the 95% prediction intervals). All model results are from simulations using 311 samples from the parameter posterior distributions. Times outside our period of interest are shaded. Lower panels: Estimated health costs and monthly scaled GDP for England for the time period of interest (1st March to 30th November 2021) [31]. The estimated health costs are based on one QALY valued at £30,000; mean values and 95% prediction intervals are shown.

### 3.1 Health outcomes

The health outcomes for the alternative roadmap scenarios are shown in Figure 3 which indicates peaks in both daily hospital admissions and daily deaths for the faster timelines that far exceed those for the default roadmap. This leads to levels of maximum hospital occupancy that exceed both the levels seen for the first wave of the virus in March 2020 (Wildtype peak occupancy 18,921) and for the Alpha peak in January 2021 (peak occupancy 31,459). For example the *Rapid* scenario generates a peak occupancy level of 41,954 (95% PI: 30,490 – 57,332) and the *One week quicker ND* scenario has a peak hospital occupancy of 30,965 (95% PI: 21,131 – 42,928). The mean values for both of these scenarios lie above or close to the level of the Alpha peak and have lower prediction interval values above the Wildtype peak (Figure 3 lower panel). This suggests that these two fastest relaxation scenarios would fail Test (3) of the roadmap conditions, as they risk a level of hospitalisation that would put overwhelming pressure on the NHS.

**Figure 3:**
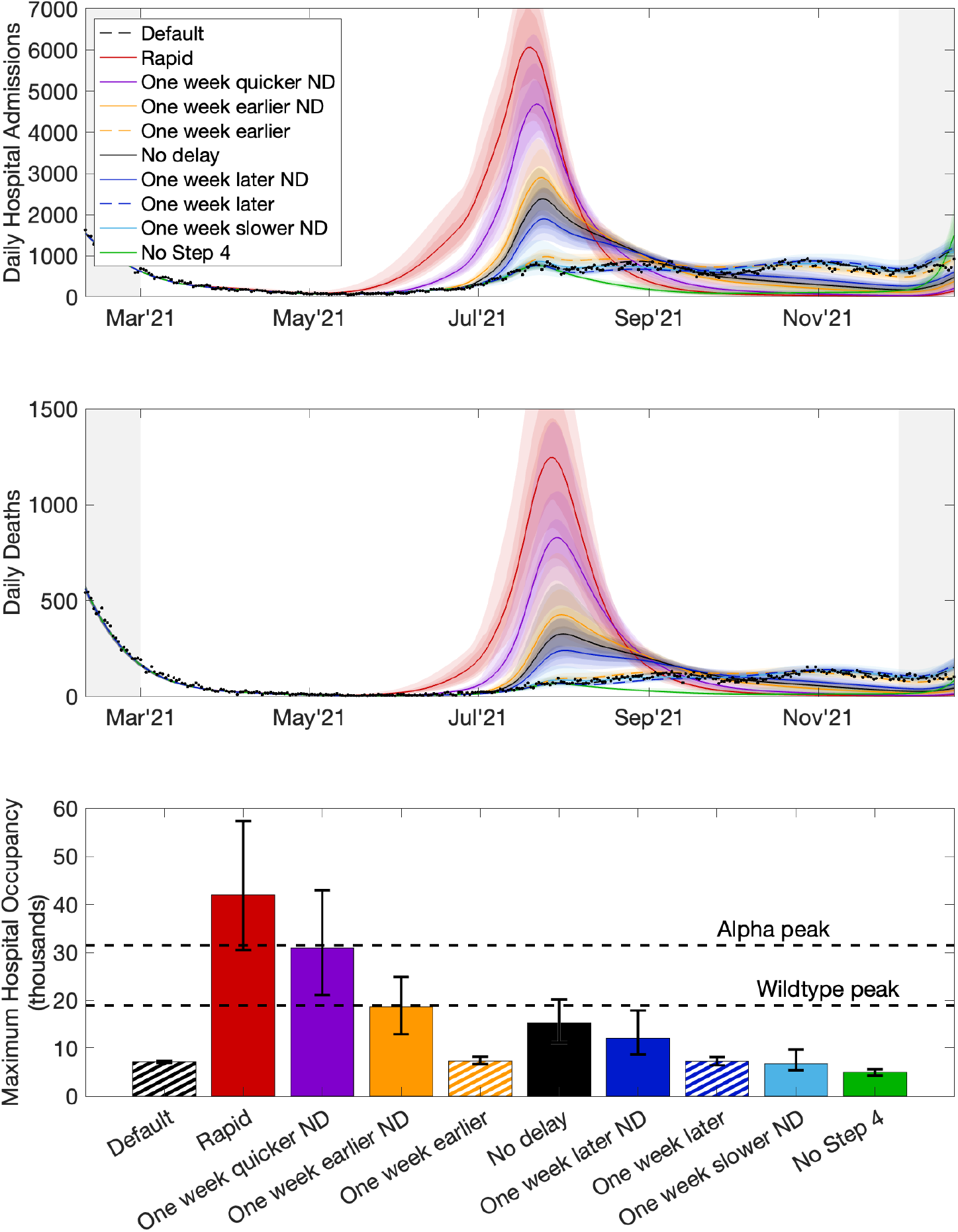
Health outcomes for range of roadmap scenarios. Upper and central panel: Daily hospital admissions and deaths for all the scenarios over the period of interest (1st March 2021- 30th November 2021) Black dots show the observed daily hospital admissions and deaths. Lower panel: Maximum hospital occupancy for all scenarios showing mean values and prediction intervals and the levels of maximum occupancy seen during the first wave in March 2020 (Wildtype peak) and during the Alpha wave in January 2021 (Alpha peak). Mean values are shown by lines in the top two panels and bars in the lower panel, with 95% prediction intervals as shaded regions in the top two panels and error bars in the lower panel.

The differences in total admissions and deaths for all alternative roadmaps compared to the default are given in Table 1. Here the general trend is for faster relaxation scenarios to lead to higher levels of hospital admissions and deaths compared to the default case. The uncertainty around the emergence of the Delta variant led to a delay in final step (pushing it from 21st June to 19th July 2021); the difference between the *Default* and *No delay* scenarios shows that this decision led to to 22,588 (95% PI: 14,047 – 36,240) fewer hospital admissions and 5,928 (95% PI: 3,714 – 12,556) fewer deaths. Other scenarios (*One week earlier* and *One week later*) allow a similar comparison and highlight the general benefit from a delay at this time. Only two scenarios lead to mean values of hospital admission and death being less than the default case – the *One week later* and *No step 4* scenarios. *One week later* is not a statistically significant improvement, with 95% prediction intervals that contain zero. Associated with these changes in hospital admissions and deaths, we predict similar changes in the number of QALYs lost across scenarios. Compared to the default, the *Rapid* roadmap has 225,870 (95% PI: 135,010 – 386,257) additional QALYs lost, the *No delay* roadmap has 43,807 (95% PI: 26,702 – 87,545) additional QALYs lost, while the *No Step 4* scenario saves 78,181 (95% PI: 57,507 – 89,202) QALYs; this impact is consistent across all adult age groups (as shown in the Supplementary Material).

**Table 1:** Difference in total hospital admissions and total deaths over the time period of interest (1st March 2021 – 30th November 2021) for each of the alternative scenarios compared to the default scenario. Values are given as mean (and 95% prediction intervals).

| Scenario | Difference in hospital admissions |  | Difference in deaths |  |
| --- | --- | --- | --- | --- |
| *Rapid* | 117,927 | (77,589 – 177,468) | 30,357 | (17,479 – 54,824) |
| *One week quicker ND* | 66,479 | (43,500 – 104,790) | 17,588 | (10,206 – 34,331) |
| *One week earlier ND* | 30,207 | (18,923 – 47,742) | 7,977 | (4,879 – 16,295) |
| *One week earlier* | 6,173 | (2,265 – 11,642) | 1,305 | (438 – 2,181) |
| *No delay* | 22,588 | (14,047 – 36,240) | 5,928 | (3,714 – 12,556) |
| *One week later ND* | 16,524 | (10,389 – 33,592) | 4,256 | (2,393 – 8,154) |
| *One week later* | -1,592 | (-6,847 – 2,744) | -298 | (-1,311 – 625) |
| *One week slower ND* | 650 | (-6,760 – 12,904) | 342 | (-1,136 – 2,877) |
| *No Step 4* | -68,630 | (-75,583 – -53,462) | -10,182 | (-11,366 – -7,300) |

### 3.2 Costs

Total health costs for each alternative strategy relative to the default roadmap are compared in Figure 4 (upper panel) for one QALY valued at £30,000. (Numerical values for the total health costs at both £30,000 and £70,000 per QALY are given in the Supplementary material.) The total cost difference is dominated by the cost of QALY losses due to death (mid-grey bar) in all scenarios and reflects the impact of the relaxation speed on severe health outcomes. Health costs for the *One week later* and *One week slower ND* scenarios are not significantly different from the default. The delay to the final step of the roadmap is estimated to have saved £1.48bn (95% PI: £0.96bn – £2.86bn) at £30,000 per QALY, or £3.24bn (95% PI: £2.09bn – £6.30bn) at £70,000 per QALY, in total health costs. For the *Rapid* roadmap the health cost difference of £7.68bn (95% PI: £4.70bn – £12.92bn) at £30,000 per QALY, or £16.71bn (95% PI: £10.16bn – £28.29bn) at £70,000 per QALY, represents more than a doubling of health costs over this period. The *No Step 4* scenario is associated with a total health cost saving of £2.89bn (95% PI: £2.16 – £3.18bn) at £30,000 per QALY, or £6.01bn (95% PI: £4.47bn – £6.67bn) at £70,000 per QALY, approximately halving the health costs.

**Figure 4:**
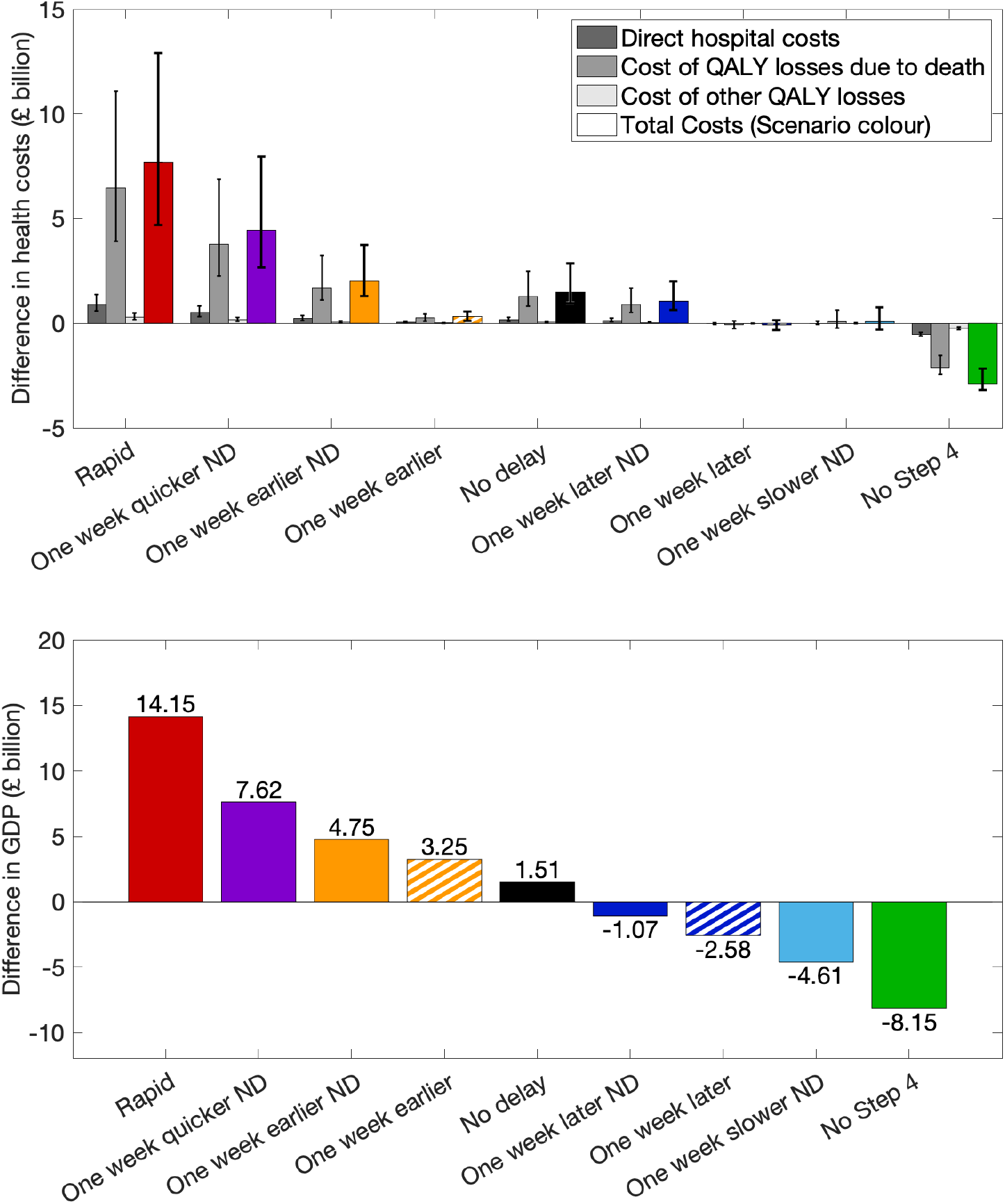
Difference in health costs and GDP for each scenario compared to the default. Upper panel: Costs are broken down into direct hospital costs, cost of QALY losses due to death, and cost due to other QALY losses (symptomatic infection, hospitalisation and Long COVID), valued at £30,000 per QALY, together with the total costs (coloured bars). Bars show mean values and error bars indicate the 95% prediction intervals for each cost. Lower panel: Estimated difference in GDP for the alternative roadmap scenarios compared to the default; this is a deterministic calculation so no prediction intervals are shown.

The pace of reopening is also reflected in the total estimated GDP over the nine months of interest; the difference in total GDP compared to the default for each of the alternative roadmaps is shown in Figure 4 (lower panel). The *Rapid* roadmap is estimated to lead to a gain in GDP of £14.15bn over the default roadmap, this is reduced to a £1.51bn gain for the *No delay* scenario, and a loss of £8.15bn as a result of remaining in Step 3 (*No Step 4*). It should be noted however that even the gains accrued by the *Rapid* roadmap, only amount to a 1% change in the total GDP over this period.

When the GDP figures are combined with the total health costs we can compare the full impact of the alternative scenarios in terms of total benefit or deficit (monetary gain or loss), see Figure 5 (and the Supplementary Material for numerical values). When one QALY is valued at £30,000 the faster (slower) relaxation roadmaps all generate a net benefit (deficit), although we have already shown that the fastest two strategies may fail Test (3) due to the high hospital occupancy burden. Not implementing the delay to Step 4 was monetarily neutral with a mean difference of just £0.03bn and prediction intervals that span zero. This NICE threshold of £30,000 per QALY [28], is used within the health service and is a reflection of the NHS’s actual spending on cost-effective treatments. For broader spending decisions, HM Treasury [29] suggests the use of £70,000 per QALY, which may be more representative of the decisions about social control measures. At this higher QALY value, only *One week earlier* consistently outperforms the default scenario, while all except this scenario and *One week earlier ND* have means that are less than the default. We note that the most rapid roadmaps are associated with larger variability in their total benefit compared to the default; this is attributable to the large outbreak sizes where small uncertainties in mixing and hence small uncertainties in the exponential growth rate can lead to large uncertainties in the magnitude of the wave.

**Figure 5:**
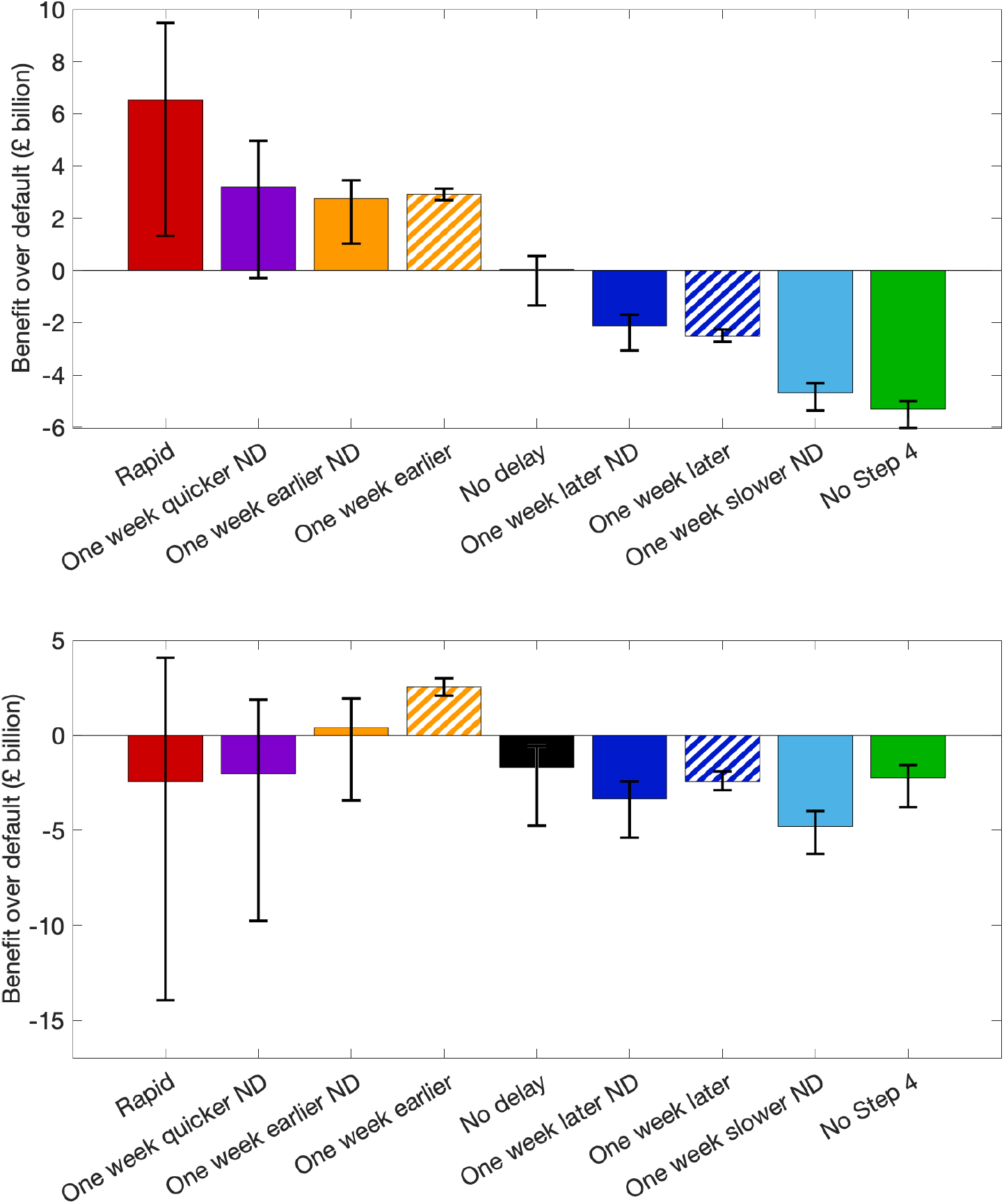
Combined economic and health benefit of alternative scenarios compared to the default roadmap over the period of interest. Upper panel: Total benefit (GDP minus health costs) compared to default with one QALY valued at £30,000. Lower panel: with one QALY valued at £70,000. Bars show mean values and error bars show the 95% prediction intervals.

Extending the analysis to a range of willingness to pay values for a QALY we calculate the proportion of simulations for each value of a QALY where there is a net benefit from the alternative scenario compared to the default roadmap. Figure 6 shows these results when the value of a QALY ranges from £10,000 to £100,000. We have calculated the percentage of parameter sets (out of 311 parameter posterior distributions) where there is a net monetary gain (GDP minus health costs) greater than for the default roadmap. We note that none of the slower relaxation roadmaps (except *No Step 4*) ever give rise to a benefit over the default case for the range of QALY values explored. The *No Step 4* strategy only produces a net gain when a QALY is valued above £90,000. All faster scenarios have a large proportion of parameter sets where there is net benefit for a QALY valued at £30,000 or less, but the proportion of sets where this occurs decreases as the willingness to pay value for a QALY increases. At £70,000 per QALY the *Rapid* and *One week quicker ND* roadmaps only have 24% and 20% of simulations that produce a net benefit. The *One week earlier* scenarios (*One week earlier* and *One week earlier ND*) give rise to the most consistent net gain across all values of a QALY, with the *One week earlier* (with the Delta delay implemented, shown as striped orange in Figure 6) always having 100% of simulations producing a net benefit compared to the default. The *No delay* strategy (when Step 4 occurs on the 21st June 2021) is only consistently beneficial for a QALY valued at £20,000 or below.

**Figure 6:**
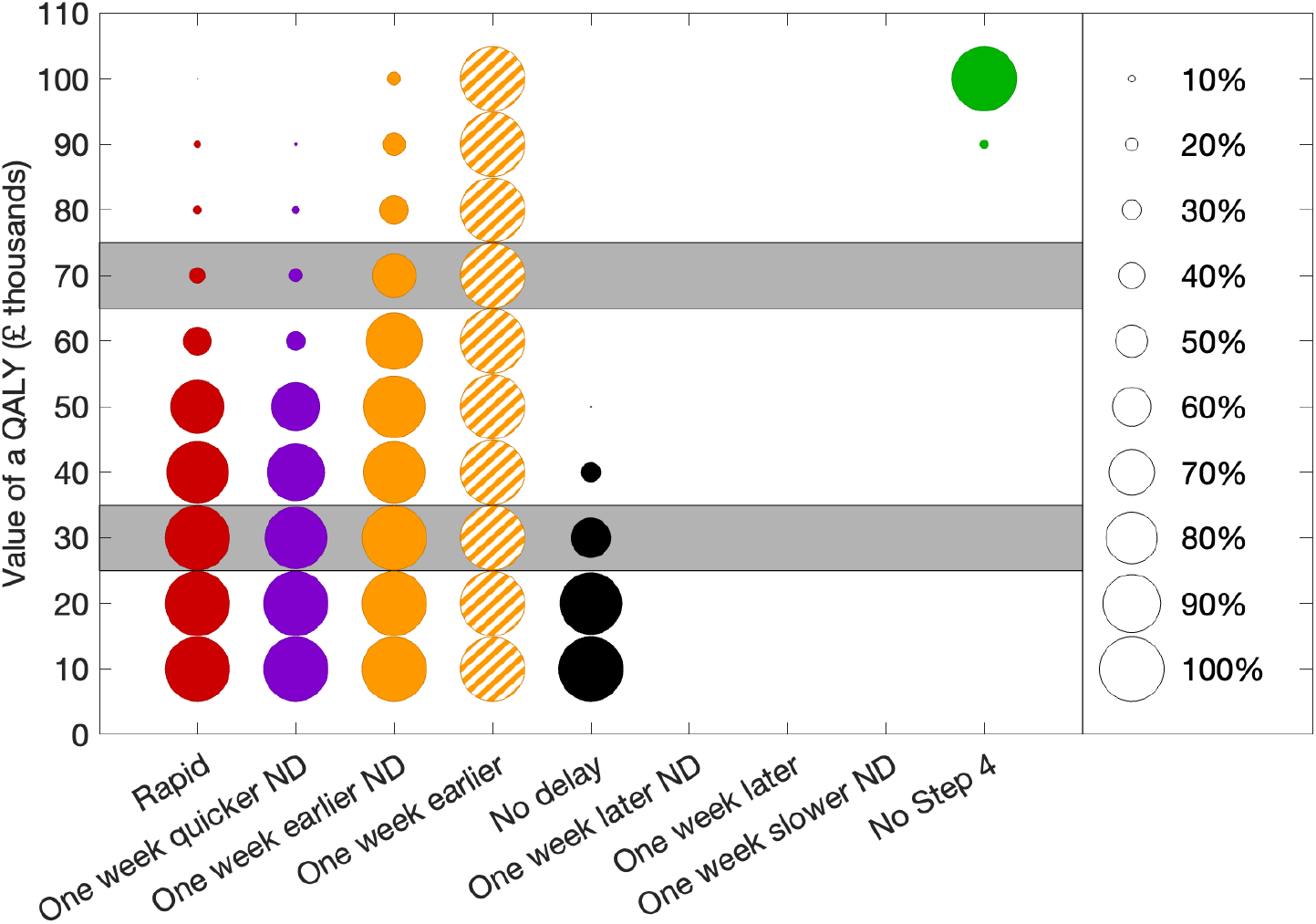
Likelihood of a net benefit over the default roadmap for each scenario across a range of QALY values. The radius of the circles indicate the percentage of model parameter sets (out of 311) where (GDP minus health costs) is greater than the default. Grey bands highlight the NICE recommended value for a QALY (£30,000 [28]) and the value used in the Treasury Green Book (£70,000 [29]). Circles on the right indicate how the radius of the circles are proportional to the percentage of parameters sets where there is a net benefit.

## 4 Discussion

A key political decision in 2021 was how best to ease COVID-19 restrictions following the third UK lockdown in January 2021 alongside the rollout of an ambitious vaccination program. The Government wanted the relaxation of restrictions to be irreversible, such that none of the step changes should place the health service under an unmanageable burden, which might require further restrictions. The role of academic-led epidemiological modelling was therefore crucial in assessing the feasibility of the chosen approach, first analysing the impact of the most recent relaxation steps and secondly predicting the likely impact of future steps. As such epidemiological modelling was important in fulfilling the “data not dates” mandate, and ensuring that the UK could reopen safely without adverse consequences. Here we examine this time period, and consider if alternative roadmaps to reopen society and the economy would have been preferable. The other major epidemiological decisions that impacted the pandemic in 2021 was the prioritisation and roll-out of vaccination; the same health-economic framework is applied to alternative vaccination scenarios in the Supplementary Material

The default roadmap [2] was derived from a purely qualitative and subjective approach for balancing the pressures of reopening the economy while not placing too high a burden on health services. It arose from a political appraisal of the benefits and harms. Yet, despite this, it performs remarkably well in our rigorous quantitative assessment. We compared the enacted roadmap against strategies where each step was taken quicker (4 weeks between each step rather than 5) or slower (6 weeks between steps), where the entire roadmap started one week earlier or later, as well as far more rapid or cautious extremes. When the Step 4 decision occurs in June 2021, corresponding to the start of the Delta wave, we also consider scenarios with and without an additional 4-week delay.

One view proposed at the time was – with vulnerable groups likely to have been vaccinated by the end of April, the Government should focus on a quick resumption of economic activity and relax all social restrictions by this point. The academic models, including the one from Warwick, warned that such a rapid opening would risk another wave of infection with severe outcomes that could put health services under strain [4]. This helped to shape the published roadmap, where the government decided on a slower relaxation of controls, with an emphasis on maintaining relatively low hospital occupancy. Our model-based investigation into alternative roadmap timelines demonstrated that a rapid relaxation of restrictions could indeed have led to a surge in severe outcomes; in particular, extremely high levels of hospital occupancy in exceedance of any observed since the start of the pandemic (Figure 3). Moreover, using our combined health-economic and economic framework we estimate that this rapid opening would not have led to a significant benefit in economic terms – a net benefit of £6.47bn (95% PI: £1.23bn – £9.45bn) at £30,000 per QALY, but a net deficit of £2.56bn (95% PI: –£3.99bn – £14.14bn) at £70,000 per QALY. It could be argued that the surge in infection (and in particular, the concern over the large number of hospital admissions and deaths) would have led to behaviour change in the public and so the severe health outcomes may not have been so pronounced. However, such a change in behaviour and the large number of unwell individuals, would likely have had a knock-on effect on the economy, from mass workplace absence and less social mixing, leading to our GDP estimates being overly optimistic.

From a pure public health perspective, remaining under restrictions for a longer period of time (*No Step 4*) is the most beneficial approach, which we estimate would have saved approximately 10,200 (95% PI: 7,300 – 11,400) lives and 68,600 (95% PI: 53,500 – 75,600) hospital admissions. However, our health-economic and economic framework suggests that the expected combined benefit of this approach is only positive if a QALY is valued at £90,000 or higher. Such a prolonged period under restrictions is also likely to lead to social problems and run the risk of a surge in cases into the winter period due to a build-up of susceptibility within the population.

Another key decision made during 2021 was the deviation from the original published roadmap timeline by imposing a delay to the final relaxation Step 4. In June, following the emergence of the Delta variant, there was considerable uncertainty about how transmissible this new variant was and how effective the vaccines would be [32, 33]. Based on sensitivity analyses around these uncertainties [8] it was decided to opt for a four week delay to the final relaxation step – in alignment with the “data not dates” approach. In terms of purely health outcomes our retrospective modelling suggests that this delay saved approximately 5,900 (95% PI: 3,7000 – 12,600) lives and 22,600 (95% PI: 14,000 – 36,200) hospital admissions. In the economic framework, assuming a QALY valued at £30,000 the decision was almost neutral; without the delay leading to a net deficit of £0.03bn (95% PI: £1.35bn deficit – £0.55bn benefit) and with a QALY valued at £70,000 estimated to produce a deficit of £1.73bn (95% PI: £0.58bn – £4.79bn) compared to the implemented roadmap.

The optimal strategy suggested by our calculations is when the entire roadmap process was begun one week earlier, such that each step (including the delay to Step 4) occurs just one week before its realised date. Given the time needed assess the impact of the January lockdown and mass vaccination, and to design a robust roadmap, it is unlikely that this faster initialisation could have been achieved.

As with any modelling venture, there are a number of approximations and assumptions that have been made. We have implicitly assumed that Gross Domestic Product (GDP) is a good characterisation of the economic health of the nation; while healthcare costs and QALY losses completely capture the nation’s health status. Both are simplifications of reality. Factors such as employment rates, inflation, and the Human Development Index are also part of any comprehensive assessment of economic prosperity. Similarly, our characterisation of health has ignored the impact of both controls and disease on mental health or on delayed treatment for other conditions. However, we believe the advantages of having a simple single metric for both health and the economy outweigh the disadvantages. A second major assumption is that the average GDP within any step and the social mixing within any step are not impacted by the timing or duration of that step – with these averages based on the observed situation in 2021. We believe this assumption is likely to hold for strategies that are a small perturbation to the default roadmap. For relaxation strategies that generate large outbreaks with many individuals requiring hospital treatment (for example, *Rapid* and *One week quicker ND*), it is plausible that changes in behaviour and also economic productivity would have occurred, but there is no agreed framework for imputing these changes which will depend on many aspects of the unfolding epidemic [34, 35].

We have also assumed that the 2020 UEFA European Football Championships (11 June to 11 July 2021) and the pingdemic (late July to early-August 2021 [36]), would have occurred at the same dates and had the same impact across all roadmap strategies. More rapid relaxations could have led to an earlier pingdemic as cases began to rise, while slower strategies (such as *No Step 4*) may have diminished the scale of the App-driven tracing. Similarly, we assume that the vaccine roll-out and uptake would be unaffected by changes to the relaxation of controls or the underlying patterns of infection. With the strategies that are the furthest from the realised roadmap (*Rapid* and *No Step 4*), it is also more likely that the population’s precautionary behaviour would be different, driven by more (or less) concern about the levels of severe disease – affecting both the epidemiology and economics. Hence, while small perturbations to the roadmap are likely to be robust, the more extreme strategies are subject to more uncertainty.

Overall, the implemented roadmap appears to have been well judged and, under the conditions imposed, appears to have balanced the health and economic burdens successfully. However, this success may have been fortuitous; future outbreaks would benefit from a quantitative modelling framework that seeks to combine the range of health, social and economic outcomes. Such projections are challenging, especially in terms of human response to changing regulations and perceived risk, but have the benefit of robustly reflecting the tradeoff between different consequences.

## Data Availability

Data from the CHESS and SARI databases were supplied after anonymisation under strict data protection protocols agreed between the University of Warwick and Public Health England. The ethics of the use of these data for these purposes was agreed by Public Health England with the Governments SPI-M(O) / SAGE committees.

## Acknowledgements

A.J.C. was funded by the University of Warwick REF impact support fund. M.J.K. was supported through the JUNIPER partnership [grant number MR/X018598/1] and by the National Institute for Health Research (NIHR) [Policy Research Programme, Mathematical and Economic Modelling for Vaccination and Immunisation Evaluation, and Emergency Response 3; NIHR204667]. The funders did not play any role in the study design, data collection and analysis, decision to publish, or preparation of the manuscript.

