## Supplementary Material for "A Health Economic Assessment of Relaxation of COVID-19 Control in England 2021"

Here we provide more details on multiple elements of the approach underpinning this work. Section 1 gives a synopsis of the model structure used to generate the alternative epidemiological scenarios. Section 2 provides additional information on the health economic framework including direct hospital costs and costs associated with QALY losses. Section 3 provides similar details on the economic framework used to predict the GDP over the time period of interest. Section 4 contains a series of additional results, as figures and tables, to supplement those in the main text. Finally, Section 5 performs a similar health economic calculation for two alternative vaccination strategies, while keeping with the default roadmap timings.

#### 1 Transmission model

For all the simulations we utilise an age-structured SEIR-type model that was employed throughout the COVID-19 pandemic, and fed into the SPI-M-O projections [1–6]. The model has been extensively documented in a number of publications: from its initial inception [7]; to its use in optimising vaccine deployment [8, 9]; to helping to shape the relaxation of controls in England following the January 2021 lockdown [10]. For completeness we provide a general overview of the model structure and fitting procedure.

The model is built around the traditional deterministic SEIR (Susceptible, Exposed, Infectious, Recovered) model framework [11], with three exposed classes to capture the distribution of times from infection to becoming infectious [12], and splitting the infectious group into symptomatic and asymptomatic infection. To this simple model we add additional structure to capture the effects of restricted social interaction during isolation whilst maintaining household transmission [7]. This fundamental model is then 'replicated' twenty-one times to mimic five-year age groups ($0-4$, $5-9$, . . ., 100+). The model is written as a large number of ODEs (ordinary differential equations).

This basic model was sufficient for the early waves of infection (from January to November 2020) with a single variant and without vaccination. During this early phase of the pandemic, the main driving parameter was the level of precautionary behaviour, which was inferred on a weekly basis and assumed to be slowly varying (with the exception of when new legislation was imposed) [13]. The precautionary behaviour was used to determined the level of social mixing and therefore the scale of transmission outside the household [7]. We also inferred a number of other parameters (including case-hospitalisation and case-mortality ratios, age-dependent susceptibility and probability of symptoms, and the relative strength of asymptomatic compared to symptomatic transmission). Our precision in estimating each of these parameters increased throughout the pandemic, as more data were accumulated; in particular the REACT2 study was vital in determining the true prevalence of infection [14].

From the age-structured symptomatically infected class, we can calculate the number of severe outcomes (hospital admissions, intensive care unit admissions and deaths), which are key public health observables and measures of concern for this pandemic - although these quantities do not impact the transmission dynamics. Our fitting is performed in a Bayesian framework, matching the data on the daily hospital admissions, hospital occupancy, ICU occupancy, deaths and proportion of community (Pillar 2) tests that are positive in each of the seven National Health Service (NHS) regions of England assuming a Poisson distribution with a mean given by the ODE model. This fitting was performed sequentially throughout the pandemic as more data became available [13], with the precautionary behaviour being the key time-varying parameter that drives the rate of change of infection.

From late 2020, and therefore of more interest to this work, variants and vaccination increased the dimensionality of our model. Each new variant (Alpha, Delta and Omicron) required a duplicate of all the infected model classes, to capture differences in transmission, risks of severe outcomes, risks of reinfection and differences in vaccine protection. The rise of each variant was captured by additionally fitting to the proportion of S-gene target failures (a proxy measure of variant-type) from TaqPath PCR testing [15, 16]. The models used in this work capture wildtype, Alpha, Delta and Omicron variants, as the main variants that affected the UK; but only the dynamics of Alpha and Delta are important for the time period under consideration.

The start of the vaccination campaign in December 2020 necessitated a further partitioning of the population by vaccination status (unvaccinated, vaccinated with first dose, vaccinated with second dose and boosted), allowing us to capture both the reduced risk of infection and the reduced risk of severe outcomes with vaccination. By late 2021, and certainly by the time Omicron invaded the UK, waning levels of protection both in terms of vaccine-induced and infection-induced immunity were added to the model in addition to booster vaccinations.

Only one modification has been made to the model code since our retrospective epidemiological investigations of the roadmap [10] and vaccine deployment [9]. In the original models, the invasion of new variants was simulated by converting a fixed proportion of current infections to the new variant (e.g. changing 1% of Alpha infections to Delta infections). This approach works well when modelling relatively small perturbations, such that there is always a similar level of infection at the point of invasion. However, given we are now looking at wide deviation in our counterfactual scenarios, we now insist on a fixed number of new variant infections entering the country. This is a more realistic assumption, although does not change the dynamics for our default models (ie at the default parameters the number and proportional approaches are identical).

#### 1.1 Precautionary Behaviour

Precautionary behaviour, termed $\phi(t)$ in our model framework [7], is conceptualised as the level of epidemiological awareness in the population that reduces risky behaviour and hence reduces transmission. Precautionary behaviour is scaled between zero (pre-pandemic behaviour) and one (only essential mixing in the population). Although we explicitly translate precautionary behaviour into a reduction in the population level mixing (between susceptible and infectious individuals), it can also account for a reduction in 'mixing' due to test, trace and isolation, mask-use or other measures that reduce the risk associated with a contact.

The infection dynamics within our model are driven by four factors: the gradual depletion of susceptibles by infection or through vaccination; the slow waning of immunity; the invasion of new variants with higher transmission; and the fluctuations in precautionary behaviour either in response to legislation or as a reaction to perceived risk. Of these,

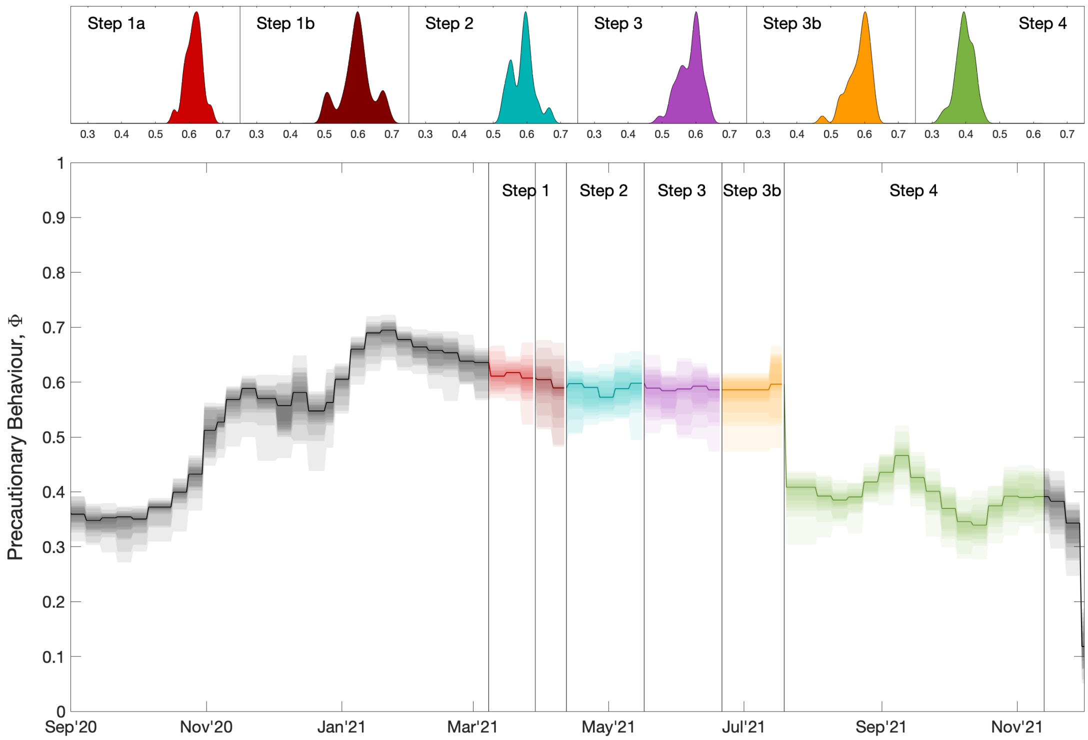


Figure S1: Inferred precautionary behaviour, $\Phi(t)$, with the impact of the Euro2020 tournament and the subsequent pingdemic removed. The distribution of mean values across each Roadmap Step are shown in the top panels.

precautionary behaviour has the potential to act over the shortest timescales, explaining the success of social distancing measures (such as lockdowns) for controlling the infection [17]. When considering alternative roadmaps for the relaxation of controls, we are considering different changes in precautionary behaviour over time. While there is some evidence that population-level responses to social distancing measures change over time as the individuals adapt to these constraints, throughout we have made the simplifying assumption that precautionary behaviour remains constant throughout a relaxation step irrespective of its length (or the timing) of that step.

Mathematically, for posterior parameter set $i$ from the MCMC inference, we have the precautionary behaviour $\phi^i(t)$ for each time, $t$, during the pandemic (see Figure S1). We first define $\Phi(t)$, based on our estimated $\phi(t)$, but with the reduction in precautionary behaviour due to the Euro2020 football (22nd June to 12th July 2021) and the increase in precautionary behaviour due to the pingdemic (20th to 26th July 2021) removed, with values linearly extrapolated from either side of these periods. We can then define the underlying precautionary behaviour associated with each step:

$$\Phi^i_{\text{StepN}} = \underset{t \,\in\, \text{StepN}}{\text{mean}} \left(\Phi^i(t)\right)$$

These step-specific precautionary behaviour values are then used throughout the steps for each new scenario (Figure S1), with the perturbation due to the Euro2020 football and subsequent pingdemic added at the appropriate times.

##### 1.2 Vaccination

The other major component that contributed to the control of COVID-19 during 2021 was vaccination. In this supplementary material we also consider the health economics of alternative vaccination strategies (Section 5), following the same format as in the retrospective study by Keeling et al. [9]. Three strategies are examined: firstly, vaccination as it occurred during 2020 and 2021, which generally prioritised older individuals; secondly, vaccination which prioritised younger individuals; and finally, vaccination where second doses are given at 3 weeks rather than 12 - for this latter strategy, we also consider whether the shorter interval leads to reduced vaccine efficacy of the second dose [18].

The impact of vaccination is a multi-faceted parameter, with different levels of protection against infection, symptoms, hospital admission, mortality and onward transmission if infected. There are also different levels of protection against different variants (although in this study it is the efficacy against Alpha and Delta that is our main interest); there are also different levels of protection offered by first and second doses, with the second dose protection depending on the between dose interval (3 or 12 weeks). Finally, there are generally higher levels of protection associated with the Pfizer/Moderna mRNA vaccine, compared to the AstraZeneca ChAdOx vaccine. All these components are summarised in Table S1; the efficacy values at 12 weeks come from UK Health Protection Agency studies of the vaccine impact in England [19]. The lower level of protection at 3 weeks (D2*) is calculated as in Keeling et al. [9] based on the lower level of neutralising antibodies observed for shorter intervals [18].

We adopt a simple methodology to achieve the different scenarios. Using the recorded pattern of vaccinations, we compile an ordered list of first doses by age for each of the seven NHS regions (between December 2020 and November 2021), and the total number of doses (first and second) administered in each region. For the shorter dose interval, on each day we prioritise administering the second dose for those that were vaccinated three weeks ago (or more), and use any surplus in the daily capacity to administer first doses in the recorded order. When prioritising younger individuals, we first give second doses for those that have been waiting 12 weeks or more, and then give first doses in the reverse order to that recorded.

#### 2 Health economic framework

Our health economic framework is based on the estimation of two main quantities: the direct costs associated with hosptial admission, hospital stay and stay in critical care (or ICU); the health losses associated with infection, captured as a loss of Quality Adjusted Life Years (QALYs). Both of these quantities will vary with disease severity and hence with age of the infected individual.

##### 2.1 Direct hospitalisation costs

Hospital episode costs for England were obtained from the NHS 2021/22 National Cost Collection Data Publication [20] using the appropriate Healthcare Resource Group (HRG)

| | AstraZeneca | | | | | | Pfizer/Moderna | | | | | |
|---|---|---|---|---|---|---|---|---|---|---|---|---|
| **Protection** | **Alpha** | | | **Delta** | | | **Alpha** | | | **Delta** | | |
| **Against** | **D1** | **D2** | **D2*** | **D1** | **D2** | **D2*** | **D1** | **D2** | **D2*** | **D1** | **D2** | **D2*** |
| Infection | 63 | 78 | 65 | 45 | 70 | 48 | 63 | 80 | 77 | 55 | 85 | 80 |
| Symptoms | 63 | 80 | 66 | 45 | 80 | 49 | 63 | 88 | 82 | 55 | 92 | 85 |
| Hospital | 80 | 90 | 90 | 80 | 95 | 95 | 80 | 93 | 93 | 80 | 99 | 99 |
| Mortality | 80 | 95 | 95 | 80 | 98 | 98 | 80 | 97 | 97 | 80 | 98 | 98 |
| Transmission | 45 | 45 | 45 | 30 | 30 | 30 | 45 | 45 | 45 | 30 | 30 | 30 |

Table S1: Percentage protection against the Alpha and Delta variants after one dose (D1) and two doses (D2) of AstraZeneca and Pfizer/Moderna vaccines. D2 is the default level of protection after 2 doses with a separation of around 12 weeks, as estimated from English data [19]; D2* are reduced values corresponding to a dose interval of 3-4 weeks [9].

codes. This covers the period 1 Apr 2021 to 31 Mar 2022 and provides costs per Finished Consultant Episode (FCE) and costs per day in ICU.

The average number of FCEs per admission was obtained from NHS Digital, Hospital Episode Statistics (HES), England 2021-22 [21], using the diagnosis code U07.1 (COVID-19 virus identified).

The weighted average hospitalisation episode cost for a non-elective stay in 2021/22, for HRG codes DX01A, DX11A and DX21A, was used to give an average cost per admission of £5267 for adults, and £3671 for children (HRG codes DX01B, DX11B and DX21B).

For critical care the weighted average cost per day was determined to be £2144 for adults using HRG codes XC01Z-XC07Z, and £2371 for children dusing HRG codes XB01Z-XB07Z.

The model structure divides the population into twenty-one five-year age groups (0-4, 5-9, . . . , 90-95, 100+) and age-varying hospital costs can be calculated based on length of stay data for COVID-19 from Murphy et al. [22] and length of stay in critical care from the 2021 ICNARC report on COVID-19 [23]. (These average length of stay, for each age-group, are shown in Figure S2). The average daily cost, $C_H^d$, of a hospital admission is given by

$$C_H^d = \frac{C_H N_H}{\sum_a L^H(a) N^H(a)}, \tag{1}$$

where $a$ denotes age-group, $C_H$ is the average cost of an admission, $N_H$ is the total number of admissions, $L^H(a)$ is the length of stay with age and $N^H(a)$ is the number of admissions in a given age group. Daily costs are calculated separately for adults ($C_H^d = \pounds 511$) and children ($C_H^d = \pounds 524$). This then gives the age-varying admission cost as

$$C^H(a) = C_H^d L^H(a). \tag{2}$$

The age-dependent cost of critical care, $C^I(a)$, is similarly the average daily cost (£2144 for adults or £2371 for children) multiplied by length of stay:

$$C^I(a) = C_I^d L^I(a). \tag{3}$$

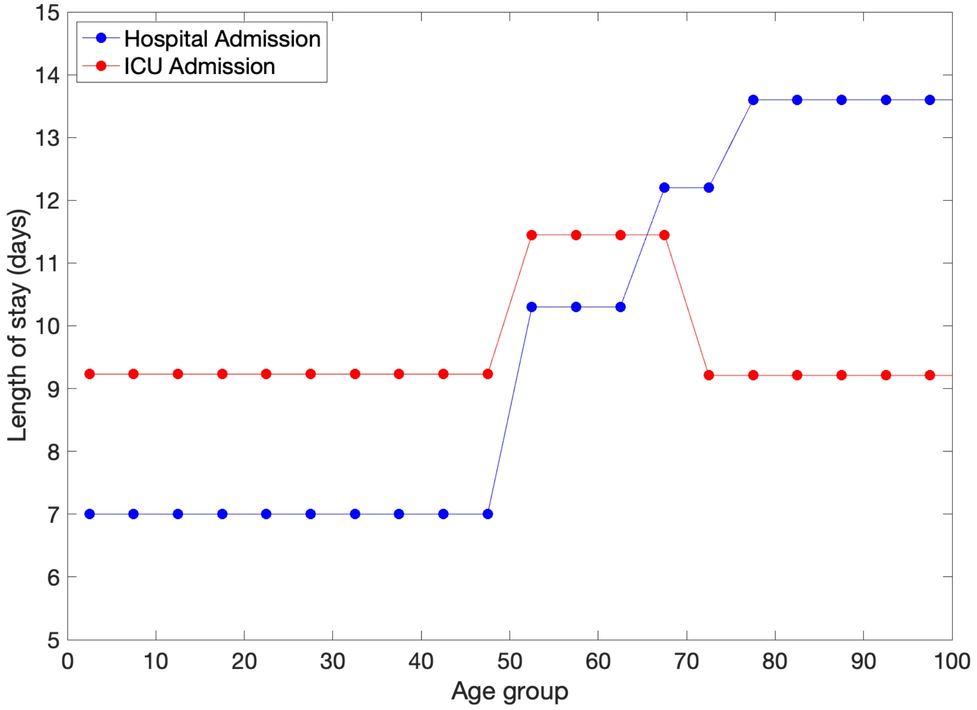


Figure S2: Length of stay with age for hospital admissions as given by Murphy et al. [22] and length of stay in ICU as given in the ICNARC report on COVID-19 [23].

#### 2.2 Cost from loss of Quality Adjusted Life Years

As well as direct hospitalisation costs there are also costs associated with quality adjusted life year (QALY) losses from symptomatic infection ($S$), hospitalisation ($H$), ICU admission ($I$), Long COVID symptoms ($L$) and costs from QALY losses due to death ($D$).

QALY losses associated with COVID-19 deaths are age-dependent and sourced from ONS [24], with adjustments for age- and sex-specific QALY population norms based on the EQ-5D-3L for the UK, and also include the UK discounting rate of 3.5% per year. The QALY losses for premature death across the twenty one age groups are shown in Figure S3.

Costs associated with QALY losses are given by

$$Cost_X = QALY \times Q_X, \tag{4}$$

where $QALY$ is the value associated with one QALY loss and $X$ is one of $S, H, I, L$ or $D$.

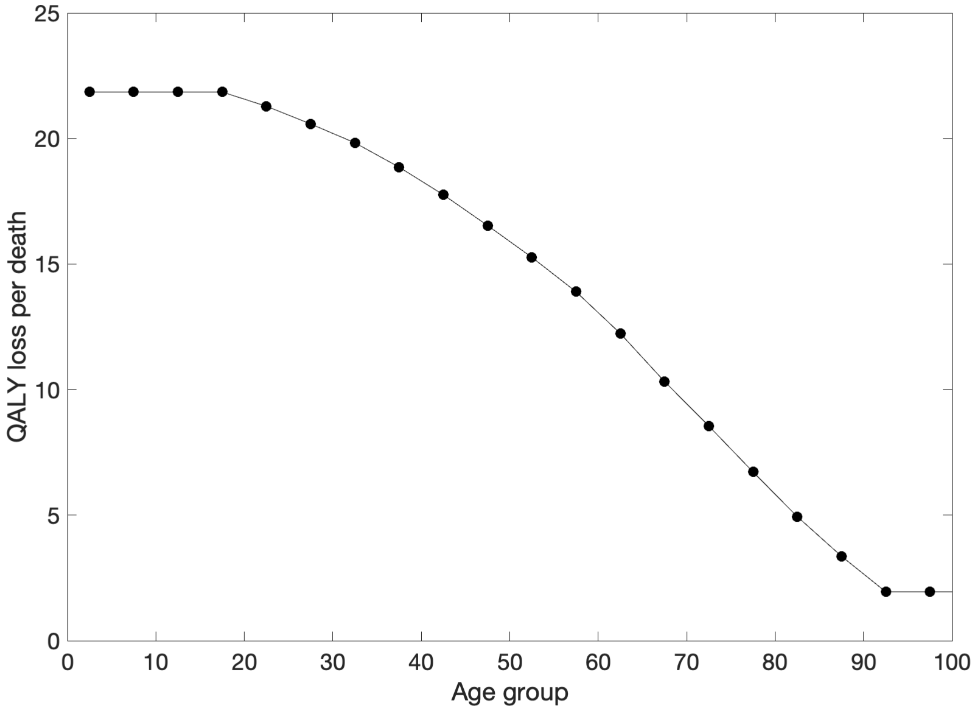


Figure S3: QALY losses from premature death across age groups, calculated as the life expectancy from a given age [24], discounted at 3.5% per year.

While QALY losses associated with death ($Q_D$) are age-dependent, the other losses are all assumed to be constant across all age groups. These other losses take the following values: QALY losses per symptomatic case, $Q_S = 0.008$; QALY losses per hospital admission, $Q_H = 0.0201$; QALY losses per ICU admission, $Q_I = 0.15$; QALY losses per episode of Long COVID, $Q_L = 0.034$, where 10% of symptomatic cases are assumed to experience Long COVID [25]. As a sensitivity analysis, we also considered using an alternative set of QALY losses [26]: $Q_S = 0.00167, Q_H = 0.031, Q_I = 0.03457, Q_L = 0.15$; these alternative values only led to a 1% change in total health costs, as expected given that QALY losses due to death dominate the calculation.

The total cost of COVID-related health impacts can then be expressed as the sum of age-dependent ($N^H$, $N^I$ and $N^D$) and non-age-dependent ($N_S$, $N_H$, $N_I$ and $N_L$) components.

$$\text{Total Cost} = \underbrace{\sum_a C^H(a)N^H(a) + C^I(a)N^I(a)}_{\text{Direct hospital costs}} + \underbrace{QALY \times \sum_a Q_D(a)N^D(a)}_{\text{Costs due to death}} + \underbrace{QALY \times (Q_S N_S + Q_H N_H + Q_I N_I + Q_L N_L)}_{\text{Costs from other QALY losses}} \tag{5}$$

#### 3 Economic framework

The Office for National Statistics produces monthly normalised GDP values for the UK [27]. This normalisation accounts for regular monthly patterns, and is defined such that a particular reference year (2022) takes an average value of 100. We scale these values such that the total GDP for 2021 is the reported £2,409,815,000 for the UK, and then scale by 0.8852 to obtain a value for England, which can be interpolated to each day of the year. This daily GDP for England is matched to each step of the default roadmap, to generate a pattern of GDP associated with each step. In order to estimate the GDP for alternative roadmap scenarios, which involve changes to the start date and duration of these steps, the daily pattern is stretched or contracted to fill the appropriate duration. Results are then amalgamated into monthly values (Figure S4). The dip in observed GDP for July (of around 0.6%) coincides with the 'pingdemic' attributed to a surge in cases following increased mixing during the 2020 UEFA European Football Championships and the emergence of the Delta variant. This effect has been maintained for July 2021 in all scenarios. The estimated monthly GDP for England for the ten different roadmap scenarios is shown in Figure S4; all roadmaps, except *No Step 4*, start and finish with the same GDP, as they start and finish with the same controls in place.

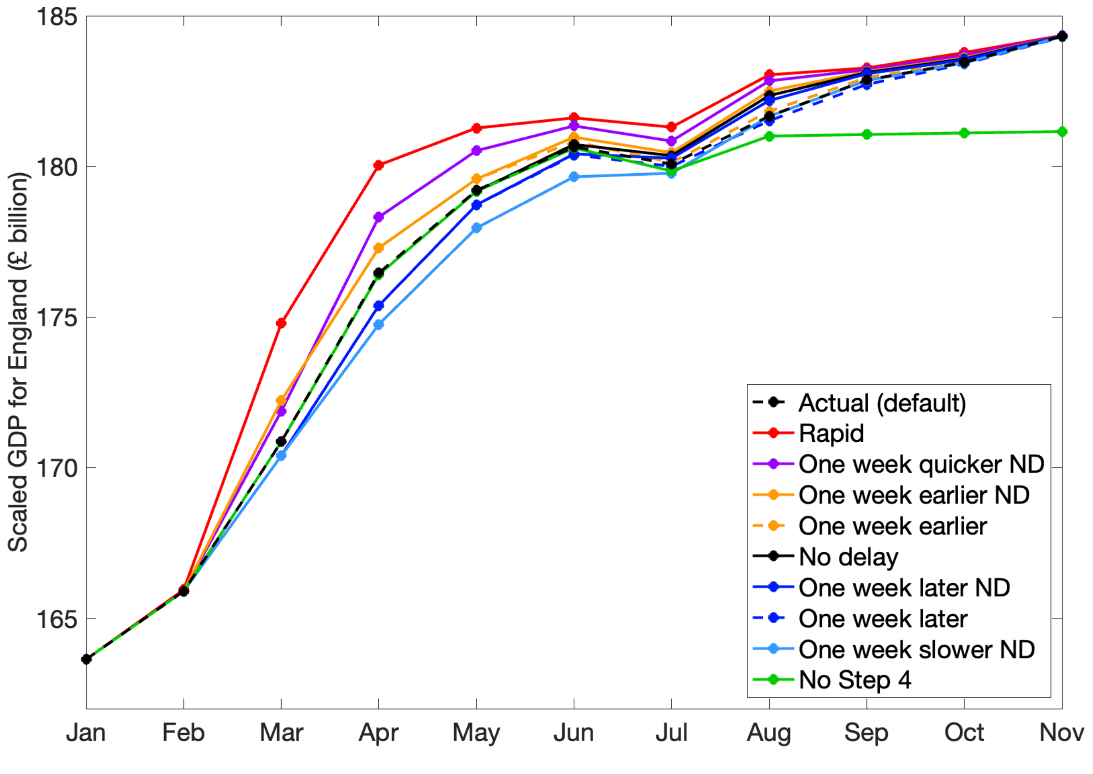


Figure S4: Assumed monthly GDP for England in 2021 for the alternative roadmaps.

### 4 Additional results

Here we present additional results of importance to the health economic assessment of the alternative roadmap scenarios compared to the default.

Associated with the changes in hospital admissions, deaths and level of infection we predict the additional QALYs lost (or gained) as a result of the alternative roadmap timelines. The total difference in QALYs lost accounts for loss of QALYs due to premature death, hospitalisation (including admission to critical care), symptomatic infection and levels of Long Covid. Figure S5 shows the difference in QALYs lost compared to the default roadmap broken down by age group and totalled across all age groups for the period of interest.

Numerical values for the total difference in health costs for each of the alternative roadmaps is given in Table S2. Costs are presented as means and 95% prediction intervals for a QALY valued at £30,000 and £70,000. The total cost difference is dominated by the cost of QALY losses due to death in all scenarios and reflects the impact of the relaxation speed on severe health outcomes. For the *Rapid* roadmap the health cost difference represents more than a doubling of health costs over this period. The *No Step 4* scenario is associated with a total health cost saving, approximately halving the health costs.

The breakdown of difference in health costs for a QALY valued at £70,000 is shown in Figure S6. This can be compared to Figure 4 of the main paper where a QALY is valued at £30,000.

| **Scenario** | **Difference in health costs at £30,000 a QALY** | | **Difference in health costs at £70,000 a QALY** | |
|---|---|---|---|---|
| *Rapid* | £7.68bn | (£4.70bn – £12.92bn) | £16.71bn | (£10.16bn – £28.29bn) |
| *One week quicker ND* | £4.45bn | (£2.67bn – £7.96bn) | £9.71bn | (£5.80bn – £17.49bn) |
| *One week earlier ND* | £2.01bn | (£1.30bn – £3.74bn) | £4.39bn | (£2.84bn – £8.24bn) |
| *One week earlier* | £0.30bn | (£0.12bn – £0.56bn) | £0.71bn | (£0.25bn – £1.19bn) |
| *No delay* | £1.48bn | (£0.96bn – £2.86bn) | £3.24bn | (£2.09bn – £6.30bn) |
| *One week later ND* | £1.06bn | (£0.63bn – £2.00bn) | £2.30bn | (£1.37bn – £4.34bn) |
| *One week later* | -£0.07bn | (-£0.32bn – £0.15bn) | -£0.15bn | (-£0.69bn – £0.31bn) |
| *One week slower ND* | £0.08bn | (-£0.30bn – £0.76bn) | £0.18bn | (-£0.64bn – £1.64bn) |
| *No Step 4* | -£2.89bn | (-£3.18bn – -£2.16bn) | -£6.01bn | (-£6.67bn – -£4.47bn) |

Table S2: Difference in total health costs over the time period of interest (1st March 2021 - 30th November 2021) for each of the alternative scenarios - given as mean (and 95% prediction intervals).

When GDP figures are combined with the total health costs we can compare the full impact of the alternative scenarios in terms of total benefit or deficit (monetary gain or loss) and numerical values are given in Table S3. When one QALY is valued at £30,000 the faster (slower) relaxation roadmaps all generate a net benefit (deficit). Not implementing the delay to Step 4 was monetarily neutral, with prediction intervals that span zero. For broader spending decisions, HM Treasury [28] suggests the use of £70,000 per QALY, which may be more representative of the decisions about social control measures.

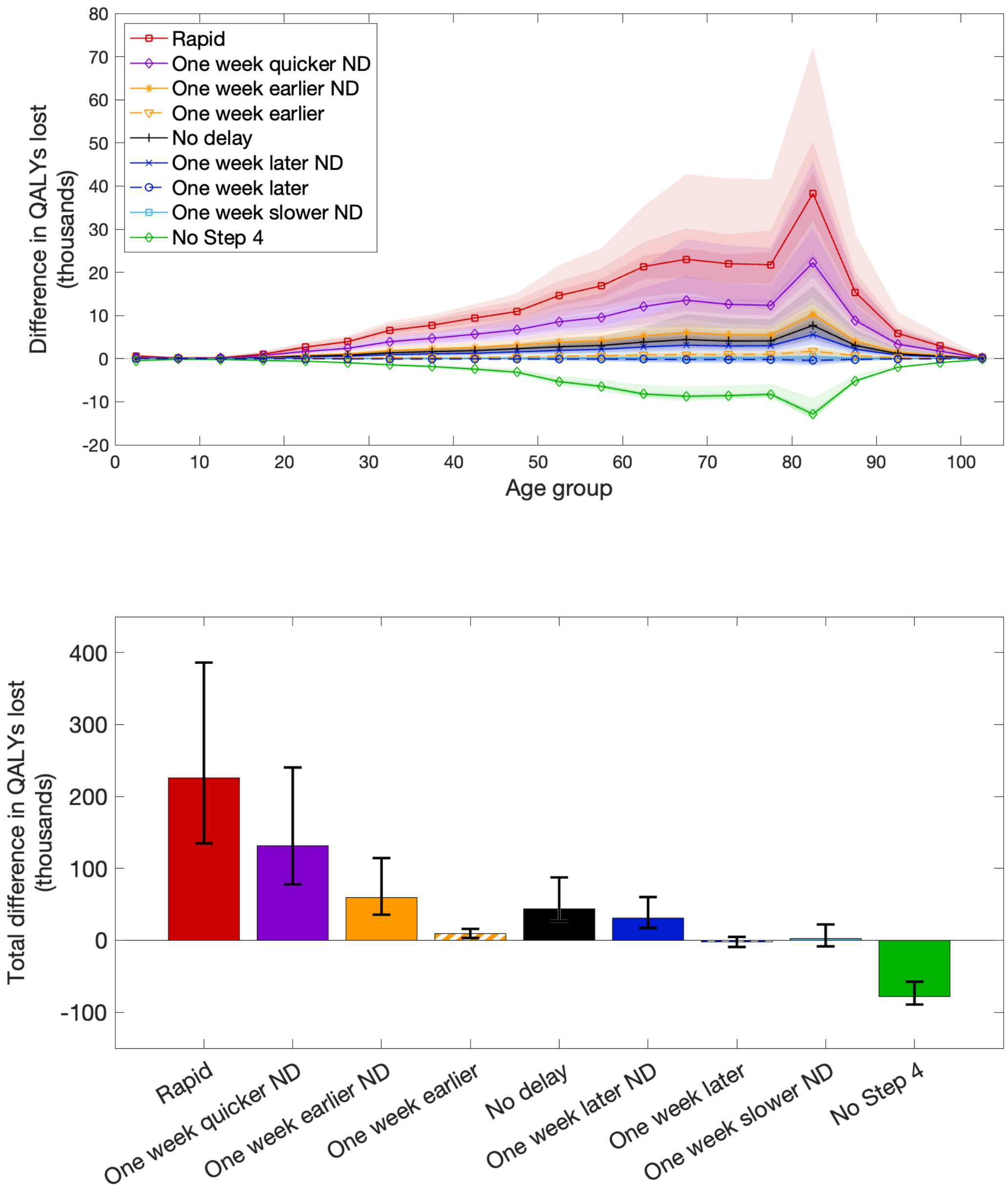


Figure S5: Loss of QALYs for each scenario relative to default showing mean (lines in upper panel and bars in lower panel) and 95% prediction intervals (shaded regions in upper panel and error bars in lower panel). Upper panel: Difference in QALY losses for each of the twenty one age groups, Lower panel: Total difference in QALY losses.

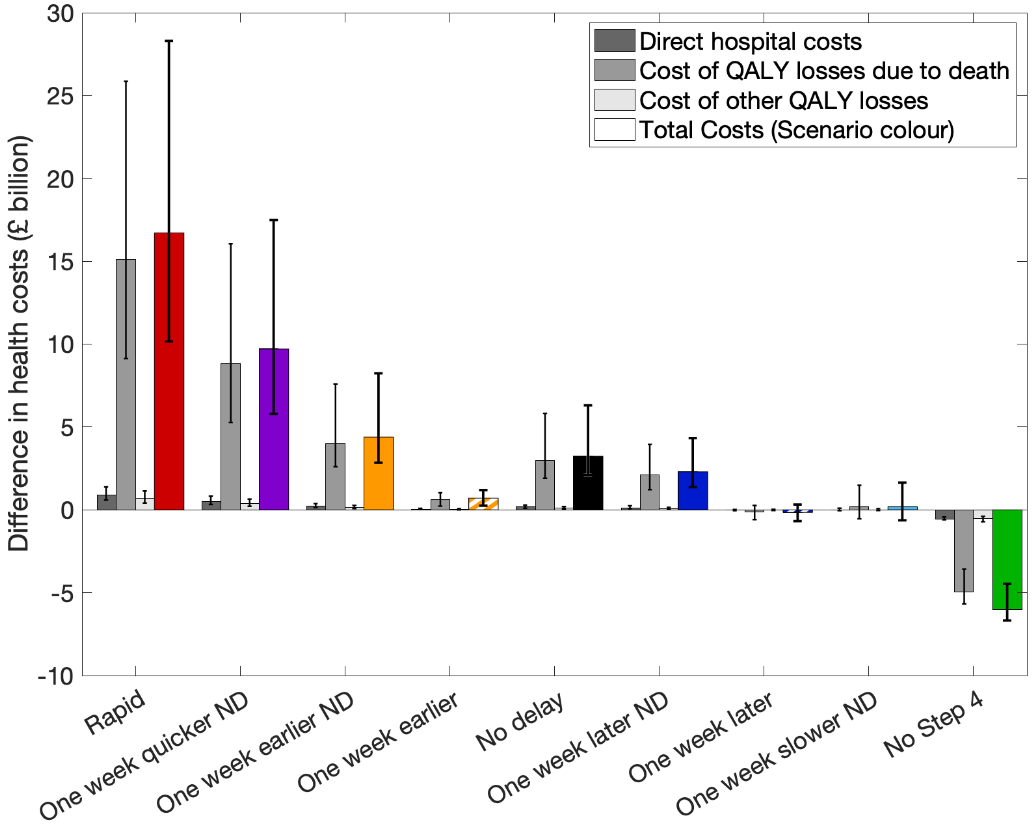


Figure S6: Difference in costs for each scenario compared to the default cost based on the cost of a QALY value of £70,000. Costs are broken down into direct hospital costs, cost of QALY losses due to death, and cost due to other QALY losses (symptomatic infection, hospitalisation and Long COVID), together with the total costs. Bars show mean values with error bars indicating the 95% prediction intervals for each cost.

| Scenario | Benefit over default at £30,000 a QALY | Benefit over default at £70,000 a QALY |
|---|---|---|
| *Rapid* | £6.47bn (£1.23bn – £9.45bn) | -£2.56bn (-£14.14bn – £3.99bn) |
| *One week quicker ND* | £3.17bn ( -£0.34bn – £4.95bn) | -£2.09bn (-£9.87bn – £1.82bn) |
| *One week earlier ND* | £2.74bn (£1.01bn – £3.45bn) | £0.36bn (-£3.49bn – £1.91bn) |
| *One week earlier* | £2.92bn (£2.69bn – £3.13bn) | £2.54bn (£2.06bn – £3.00bn) |
| *No delay* | £0.03bn (-£1.35bn – £0.55bn) | -£1.73bn (-£4.79bn – -£0.58bn) |
| *One week later ND* | -£2.13bn (-£3.07bn – -£1.70bn) | -£3.37bn (-£5.41bn – -£2.44bn) |
| *One week later* | -£2.51bn (-£2.73bn – -£2.26bn) | -£2.43bn (-2.89bn – -£1.89bn) |
| *One week slower ND* | -£4.69bn (-£5.37bn – -£4.31bn) | -£4.79bn (-£6.25bn – -£3.97bn) |
| *No Step 4* | -£5.26bn (-£6.00bn – -£4.97bn) | -£2.14bn (-£3.68bn – -£1.48bn) |

Table S3: Net benefit (GDP gains minus Health costs) over the default case for the time period of interest (1st March 2021 - 30th November 2021) for each of the alternative scenarios - given as mean (and 95% prediction intervals).

At this higher QALY value, only *One week earlier* consistently outperforms the default scenario, while all except this and *One week earlier ND* have means that are less than the default.

### 5 Extension of health economic framework to vaccine priority and dose interval

As an extension to these roadmap results it is also possible to estimate the cost and impact of other decisions taken during the UK COVID-19 vaccination campaign. The first major decision was to define the priority vaccine ordering which was based on the most vulnerable first, and generally followed an oldest and most vulnerable to youngest adult ordering [29], with the aim of preventing severe disease as opposed to reducing infection [30]. Another major decision was to increase the interval between first and second doses from three weeks to twelve weeks [31, 32]. The health implications of these decisions have been investigated previously [9] which showed that the vaccine ordering had a large impact on severe health outcomes, with the decision to prioritise the oldest (and most vulnerable) for vaccination leading to large savings in hospital admissions and deaths, compared to targeting younger individuals in an attempt to reduce transmission. In terms of changes to the dose interval, the twelve-week dose interval strategy was also estimated to have been significantly beneficial, in terms of averted hospital admissions and deaths, over the first ten months of the vaccination campaign. Using the health costs derived in this paper it is now possible to place these decisions in a health economic framework.

Three alternative dosing scenarios are considered: one where the second dose is given at 3 weeks (instead of the change to 12 weeks) where the vaccine efficacy remains unchanged; one where the dose interval is 3 weeks but with account taken of the impact this has on reducing vaccine efficacy [18]; and one where the priority ordering is reversed, so that younger individuals are the first to receive the vaccine, progressing through the age groups to the oldest individuals. If account is taken of the reduced vaccine protection under the 3 week dose interval, the levels of protection conferred by the second dose against infection and symptoms are given in Table S1.

The daily hospital admissions and deaths across the period of interest (8th December 2020 - 1st September 2021) are shown in Figure S7, and the difference in total hospital admissions and deaths for the three scenarios compared to the default model (second dose at 12 weeks) are given in Table S4.

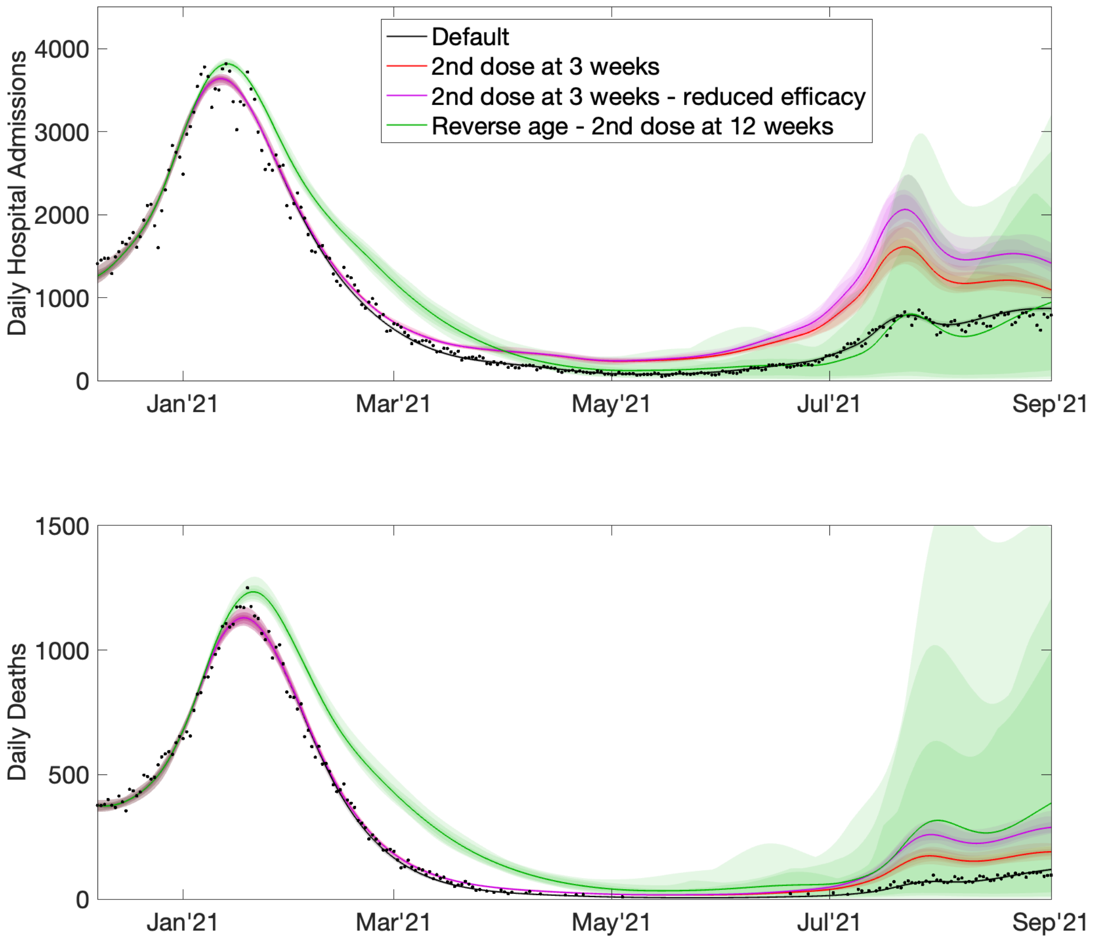


Figure S7: Health outcomes for changes to vaccine dose interval and priority ordering. Daily hospital admissions (top panel) and daily deaths (lower panel). Lines are mean values and shaded regions the 95% prediction intervals.

We evaluate costs based on NICE cost-effectiveness thresholds where a QALY is valued between £20,000 and £30,000. Default costs are £10.47bn (95% PI: £10.23bn – £10.77bn) at £20,000 per QALY or £14.61bn (95% PI: £14.28bn – £15.01bn) for a QALY valued at £30,000. We find that the enacted 12-week dose interval, compared to the counterfactual 3-week interval, saved between 57,431 and 79,683 hospital admissions and averted between 6,628 and 10,703 deaths, depending on the assumption about vaccine efficacy for

| Scenario | Difference in Hospital admissions | Difference in Deaths |
|---|---|---|
| 2nd dose at 3 weeks | 57,431 (48,944 – 71,296) | 6,628 (5,383 – 8,424) |
| 2nd dose at 3 weeks - reduced efficacy | 79,683 (63,945 – 97,739) | 10,703 (7,851 – 13,383) |
| Reverse age - 2nd dose at 12 weeks | 31,710 (-23,841 – 131,116) | 30,002 (10,821 – 82,869) |

Table S4: Difference in total hospital admissions and deaths compared to the default over period of interest (8th December 2020-1st September 2021).

the second dose (95% prediction intervals given in Table S2). This is estimated to equate to a cost saving of between £1.64bn and £2.50bn at £20,000 a QALY, or between £2.24bn and £3.44bn at £30,000 a QALY; a saving of between 13% and 19% depending on the assumptions about vaccine efficacy. (Details of savings in severe disease and costs are given in Tables S4 and S5, and Figure S8).

| Scenario | Difference in costs at £20,000 a QALY | Difference in costs at £30,000 a QALY |
|---|---|---|
| 2nd dose at 3 weeks | £1.64bn (£1.36bn – £2.15bn) | £2.24bn (£1.85bn – £2.94bn) |
| 2nd dose at 3 weeks - reduced efficacy | £2.50bn (£1.93bn – £3.27bn) | £3.44bn (£2.65bn – £4.50bn) |
| Reverse age - 2nd dose at 12 weeks | £2.43bn (£0.14bn - £7.92bn) | £3.54bn (£0.31bn - £11.44bn) |

Table S5: Difference in total costs compared to the default over the initial vaccination period (8th December 2020-1st September 2021).

Prioritising the oldest and most vulnerable first for vaccination, compared to vaccination in reverse order, is estimated to have saved 30,002 (95% PI: 10,821 – 82,869) lives and averted 31,710 (95% PI: -23,841 – 131,116) hospital admissions with an associated cost saving of £2.43bn (95% PI: £0.14bn - £7.92bn) assuming £20,000 for a QALY, and £3.54bn (95% PI: £0.31bn – £11.44bn) at £30,000 a QALY; a saving of around 19%. We note that the longer-term behaviour with vaccination in reverse order is highly uncertain - we attribute this to the waning of immunity in the younger age groups that drive the Delta wave, making the results much more sensitive to changes in social mixing in July and August 2021.

These results suggest that the JCVI's decisions taken in late 2020 with very limited data were highly beneficial in reducing the health costs associated with COVID-19 cases. These decisions were associated with the prioritisation and timing of vaccination, rather than any changes to the uptake or capacity to vaccinate; therefore they represent net savings without any additional expenditure.

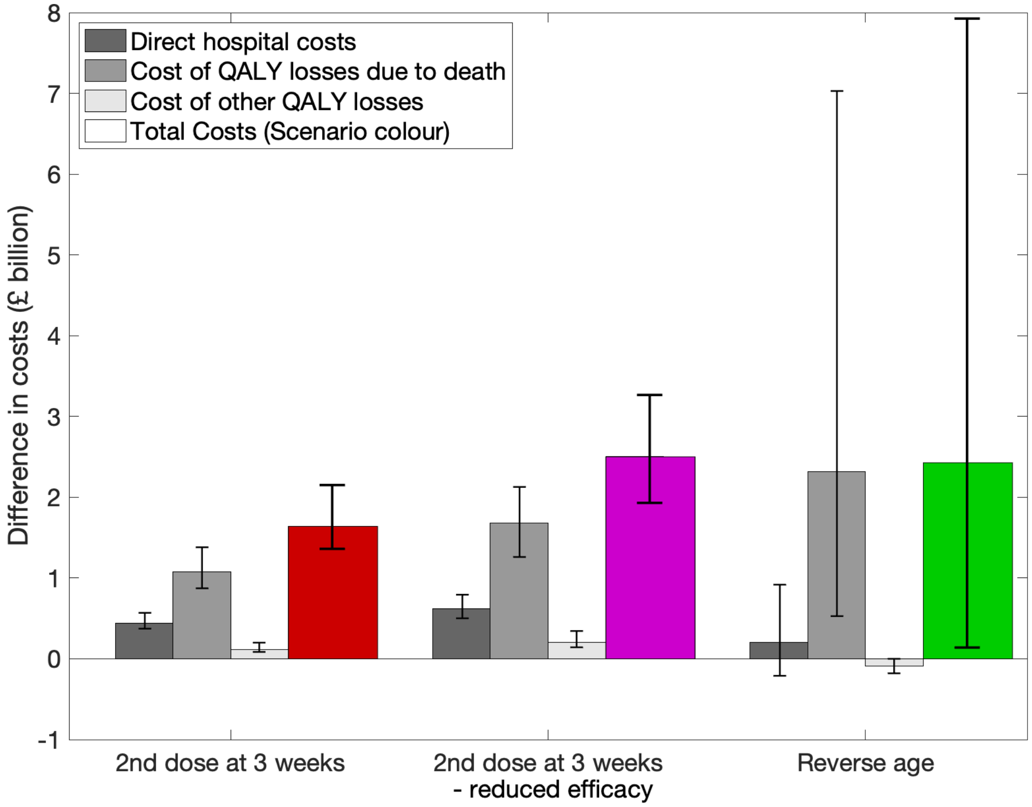


Figure S8: Breakdown of health cost differences when one QALY is valued at £20,000. Shaded bars are mean values, while error bars show the 95% prediction intervals.
